# Bayesian Borrowing of External Information in Clinical Trials: A Comparison of MAP, RMAP, and SAM Priors

**DOI:** 10.64898/2026.08.26.26360843

**Authors:** Leena Choi, Elizabeth McNeer, Cole A. Beck, Jeffrey L. Neul

## Abstract

Bayesian borrowing of external information can improve trial efficiency, particularly in pediatric and rare disease settings where patient populations are limited, but may introduce bias and inflate the Type I error rate when the trial differs from external studies. Recent U.S. Food and Drug Administration (FDA) draft Bayesian guidance emphasizes careful evaluation of external information, prior specification, and assessment of operating characteristics.

This paper compares three meta-analytic-predictive (MAP)-based methods for Bayesian borrowing: the MAP prior, robust MAP (RMAP) prior, and self-adapting mixture (SAM) prior. An adaptive platform trial design in Rett syndrome is used as a case study. Simulation studies evaluate frequentist operating characteristics under varying prior–data conflict, between-study heterogeneity, treatment effects, and clinically significant differences (CSDs) for the SAM prior.

The MAP prior achieved the greatest efficiency when external and current data were compatible but exhibited the largest bias under substantial prior–data conflict. The RMAP priors improved robustness through fixed robust-component weights, whereas the SAM prior adaptively adjusted borrowing and was less sensitive to prior–data conflict while retaining efficiency gains when the data were compatible. Although the CSD influenced the degree of adaptive borrowing, as reflected by effective sample size, it had only a modest impact on frequentist operating characteristics. Sensitivity analyses using a skeptical robust component yielded similar qualitative conclusions, while accentuating the differences between the MAP and RMAP priors.

These findings provide guidance for evaluating and selecting MAP-based borrowing strategies before trial implementation, particularly in rare disease settings, consistent with current FDA recommendations.

## 1 Introduction

Randomized clinical trials (RCTs) remain the gold standard for evaluating the efficacy and safety of new therapeutic interventions. However, conducting adequately powered RCTs may not be feasible in settings with small patient populations, such as pediatric and rare disease settings, because of limited patient availability, ethical concerns regarding placebo allocation, and recruitment constraints [3].

Recent advances in Bayesian methodology have enabled the formal incorporation of external information, including previous clinical trials, natural history studies, and real-world data, into ongoing clinical trial designs [20]. Such approaches are particularly attractive in rare disease settings, where relevant external information often exists but the number of patients available for prospective trials is limited [3].

Borrowing external information offers several potential advantages [3, 20]. It can reduce the number of participants assigned to the concurrent control arm, improve statistical efficiency, lower study costs, and shorten trial duration. It may also enhance trial feasibility and address ethical concerns associated with allocating patients to placebo or less favorable treatment groups. However, incorporating external information also presents important challenges. Differences in patient populations, study conduct, endpoint definitions, and changes in the standard of care may result in prior–data conflict, leading to biased inference and potentially compromising the operating characteristics of the trial if external information is borrowed inappropriately [3, 24].

The U.S. Food and Drug Administration (FDA) recently published draft guidance on the use of Bayesian methods in clinical trials [23]. The guidance emphasizes the careful evaluation of external information, assessment of exchangeability between external and current data, transparent prior specification, and comprehensive simulation studies to evaluate frequentist operating characteristics under plausible prior–data conflict scenarios [3]. These considerations are particularly important in pediatric and rare disease trials, where Bayesian borrowing may substantially improve trial efficiency but inappropriate borrowing may adversely affect inference.

Motivated by these regulatory considerations, this paper compares several meta-analytic-predictive (MAP)-based prior methods for borrowing external information in clinical trials. Specifically, we consider the MAP prior [12], two robust MAP (RMAP) priors with different fixed mixture weights [19], and the self-adapting mixture (SAM) prior [29], which represent different strategies for mitigating the effects of prior–data conflict when borrowing external information. An adaptive platform trial design in Rett syndrome is used as a case study to illustrate the implementation of these methods. Extensive simulation studies then compare their frequentist operating characteristics across varying degrees of prior–data conflict, between-study heterogeneity, treatment effect, and clinically significant difference for the SAM prior. The simulation framework follows the recommendations of the FDA draft guidance [23] by pre-specifying borrowing methods and evaluating their frequentist operating characteristics before trial implementation.

The remainder of this paper is organized as follows. Section 2 provides a brief overview of MAP-based prior methods for Bayesian borrowing of external information. Section 3 introduces the design of an adaptive platform trial in Rett syndrome used as a case study and reviews the evaluation of external information based on the FDA draft guidance. Section 4 presents the simulation study comparing the frequentist operating characteristics of the MAP, RMAP, and SAM priors under a range of plausible scenarios. Finally, Section 5 concludes with a discussion of the practical implications, limitations, and future directions for Bayesian borrowing in clinical trial design.

## 2 Overview of Bayesian Borrowing Methods

### Notation

Consider a new clinical trial comparing an investigational treatment with a control, and suppose that external data for the control arm from previous studies are available. Let *θ* denote a generic parameter in the statistical model.

Let *D* and *D*_0_ denote the data from the current study and the external data, respectively. Let *L*(*θ | D*) and *L*(*θ | D*_0_) denote the corresponding likelihood functions for *D* and *D*_0_, respectively, under a general statistical model. Let *π*_0_(*θ*) denote an initial prior distribution, which is typically chosen to be non-informative or weakly informative. For simplicity, throughout this paper the terms non-informative and weakly informative are used interchangeably to refer to priors that contribute minimal information relative to the likelihood.

### Meta-Analytic-Predictive (MAP) Prior

The MAP prior is constructed from a Bayesian hierarchical model that assumes exchangeability of study-specific parameters arising from a common distribution [21]. It is based on a meta-analytic framework that links study-specific parameters across the external studies. The posterior predictive distribution for a new study is then used as the prior distribution for the corresponding parameter in the current study [12].

We illustrate the construction of the MAP prior using a continuous endpoint. Let *D*_0_ = (*y*_1_*, . . ., y_H_*) denote the study-specific estimates from the external control arms, where *y_h_* denotes the observed estimate from external study *h*. Under the normal-normal hierarchical model, study-specific estimates are assumed to follow:

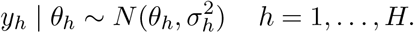

At the second level, the true study-specific parameters *θ_h_* are assumed to arise from a common normal distribution:

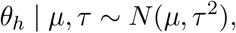

where *µ* denotes the overall mean across studies and *τ* ^2^ represents the between-study variance, which quantifies the degree of heterogeneity.

At the third level, prior distributions are specified for the hyperparameters:

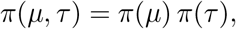

where *π*(*µ*) is typically a weakly informative prior, whereas *π*(*τ*) requires more careful specification because it reflects prior beliefs about the between-study heterogeneity and therefore influences the degree of borrowing.

Let *θ^∗^* denote the parameter for a new study. Under the hierarchical model,

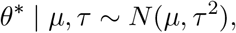

and the MAP prior is defined as the posterior predictive distribution of *θ^∗^*:

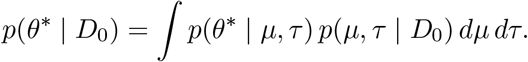

Because the MAP prior is constructed from a Bayesian hierarchical model, the degree of borrowing is determined by the estimated between-study heterogeneity rather than being specified directly. When the number of external studies is small, the between-study heterogeneity may be estimated imprecisely, making the MAP prior sensitive to the choice of the prior for the heterogeneity parameter, *π*(*τ*). Consequently, careful specification of the prior for the heterogeneity parameter is an important consideration when constructing the MAP priors. Its impact is examined in the simulation study.

### Robust MAP (RMAP) Prior

The RMAP prior extends the MAP prior approach by introducing a mixture prior. It combines two components: the MAP prior component, *π_MAP_* (*θ | D*_0_), and a robust component, *π*_0_(*θ*), which is typically specified as a vague (weakly informative) prior:

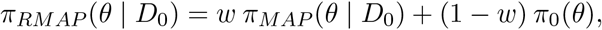

where *w* is a prespecified weight representing the prior probability that the current study is exchangeable with the external studies [19]. Following the convention used throughout this paper, the weight is assigned to the MAP prior component, whereas Schmidli et al. [19] assign the weight to the robust component. The MAP prior and RMAP priors used in the simulation study are constructed using the RBesT R package [27].

The robust component provides protection against prior–data conflict, that is, situations in which the current study data are inconsistent with the external information. By allocating a portion of the prior mass to a vague prior, the RMAP prior can reduce the influence of the external information when substantial conflict is present.

The choice of *w* plays a critical role in determining the degree of borrowing. Larger values of *w* result in greater borrowing from the external data, whereas smaller values provide greater protection against potential prior-data conflict. However, selecting an appropriate value of *w* can be challenging because the extent of agreement between the current and external studies is typically unknown at the design stage. This limitation motivates the use of dynamic borrowing approaches that adapt the degree of borrowing based on the observed data [24, 29]. One such approach, the SAM prior, is described in the next section.

### Self-Adapting Mixture (SAM) Prior

Similar to the RMAP prior, the SAM prior [29] is a mixture of two components: an informative component, *π*_1_(*θ | D*_0_), and a robust component, *π*_0_(*θ*), defined as

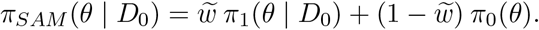

Unlike the RMAP prior, the mixture weight *w* is estimated from the current data according to the degree of similarity between the current study and the external information. Consequently, the SAM prior implements dynamic borrowing by allowing the degree of borrowing to adapt to the observed agreement between the current and external data.

To estimate the weight, the method conceptually compares two scenarios: consistency between the current and external data under *H*_0_ : *θ* = *θ*_0_, versus inconsistency under *H*_1_ : *θ* = *θ*_0_ + *δ* or *θ* = *θ*_0_ *− δ,* where *δ* is a clinically significant difference specified *a priori*, and *θ*_0_ denotes the value suggested by the external information. The relative support for these two scenarios is quantified using the likelihood ratio

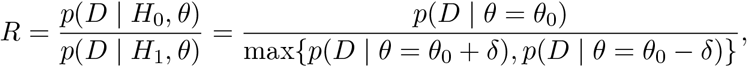

where *p*(*D | ·*) denotes the likelihood. The mixture weight *w* is then specified as a function of *R*. Following the recommendation of Yang et al. [29], we use 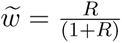. In the simulation study, the corresponding mixture weights were calculated using the SAMprior R package [28]. Smaller values of *R* indicate less agreement between the current and external data, resulting in a smaller weight assigned to the informative prior and consequently less borrowing. Conversely, larger values of *R* indicate greater agreement and therefore increased borrowing.

This approach allows borrowing to be reduced automatically in the presence of prior-data conflict, an important consideration from a regulatory perspective. Unlike the RMAP prior, the informative component *π*_1_(*θ | D*_0_) is not restricted to a MAP prior and may be any informative prior constructed from external data. In the simulation study, however, we use a MAP prior as the informative component to facilitate comparisons with other MAP-based borrowing approaches. We also investigate the impact of the choice of the clinically significant difference on the degree of borrowing and the resulting operating characteristics of the SAM prior.

## 3 Case Study: Rett Syndrome Clinical Trial

Rett syndrome is a rare neurodevelopmental disorder primarily caused by mutations in the *MECP2* gene and occurs almost exclusively in females [5], with an estimated incidence of approximately 1 in 10,000 girls by age 12 years in the United States [11]. There is currently no cure for Rett syndrome, and available treatments are limited to symptom management. Trofinetide is currently the only approved therapy for symptom improvement [14]; however, some patients are unable to tolerate treatment, highlighting a substantial unmet need for therapies with improved safety and tolerability profiles. Given the limited patient population, efficient clinical trial designs are critically needed.

As a case study, we consider a proposed phase IIa randomized, double-blind, placebo-controlled platform trial in Rett syndrome evaluating repurposed therapies, including ketamine, vorinostat, and donepezil. The estimand attributes [2] are summarized below and in Table 1.

**Table 1:** Estimand Attributes.

| Estimand Attribute | Definition |
| --- | --- |
| **Target population** | Females between 5-20 years old with Rett syndrome |
| **Treatment condition** | **Placebo arm:** Participants will receive placebo drug<br>**Active arms:** Participants will receive one of candidate drugs |
| **Endpoints** | **Endpoint 1:** Change in Caregiver Rett Syndrome Behavior Questionnaire (RSBQ) from baseline to Week 16<br>**Endpoint 2:** Clinical Global Impression of Improvement (CGI-I) at Week 16 |
| **Population-level summary** | The mean difference in the endpoints between placebo arm and an active arm |
| **Intercurrent events** | **Strategy for handling intercurrent events** |
| – Discontinuation of treatment | *Treatment policy strategy:*<br>The variables will be continuously collected, irrespective of treatment discontinuation for any reason |
| – Death<sup>a</sup> | *While-alive strategy:*<br>The variables collected up to death will be included in analysis |

<sup>a</sup> For safety endpoints, death is handled differently using a composite strategy.

The target population consists of females aged 5 to 20 years with Rett syndrome. The treatment conditions include a placebo arm and multiple active treatment arms, with participants randomized to receive either placebo or one of the candidate therapies.

Two efficacy endpoints are considered: (1) change from baseline to Week 16 in the Caregiver Rett Syndrome Behavior Questionnaire (RSBQ) score and (2) the Clinical Global Impression of Improvement (CGI-I) score at Week 16. For each endpoint, the population-level summary measure is defined as the mean difference between an active treatment arm and the placebo arm.

Intercurrent events are handled using predefined strategies. A treatment policy strategy is applied to treatment discontinuation, whereby outcome data are collected and analyzed regardless of treatment adherence. For death, which is expected to be rare in this population, a while-alive strategy is used, whereby all observed data collected prior to death are included in the analysis.

### Study Design

The proposed trial was designed within a Bayesian framework incorporating multiple active treatment arms with a shared placebo arm, interim futility monitoring, and adaptive continuation of promising treatment arms. This design allows ineffective arms to be discontinued, emerging therapies to be added, and additional participants to be allocated to treatments demonstrating early evidence of benefit. The primary objective is to identify candidate therapies suitable for evaluation in subsequent confirmatory trials.

A total of 25 participants are planned for each treatment arm, with an interim analysis planned after 13 participants per arm have completed the study to inform go/no-go decisions for each treatment.

#### Review of external information

Before constructing informative priors for the Bayesian design, we reviewed external control data from two previous randomized clinical trials to assess their suitability for Bayesian borrowing: (1) the Glaze trial [4], a phase II study evaluating the safety and tolerability of trofinetide; and (2) the Lavender trial [14], a phase III study evaluating the efficacy, safety, and tolerability of trofinetide. Both trials used the same efficacy endpoints considered in the current study: change from baseline in the RSBQ score and the CGI-I score. For the RSBQ, a negative change from baseline indicates symptom improvement, with larger negative values representing greater improvement. For the CGI-I, lower scores indicate greater improvement, with a score of 4 indicating no change. Summary statistics for the placebo arms are presented in Table 2.

**Table 2:** Summary Statistics of Efficacy Endpoints for the Control Arm.

| RSBQ |  |  |  | CGI-I |  |  |  |
| --- | --- | --- | --- | --- | --- | --- | --- |
| Study | N | Mean | SE | Study | N | Mean | SE |
| Glaze | 24 | -2.3 | 1.54 | Glaze | 24 | 3.5 | 0.14 |
| Lavender | 85 | -1.7 | 0.98 | Lavender | 86 | 3.8 | 0.06 |

RSBQ: change from baseline to Week 16 in the Caregiver Rett Syndrome Behavior Questionnaire score; CGI-I: the Clinical Global Impression of Improvement score at Week 16; SE: the standard error of the mean

Because not all external data sources are relevant to a given trial, careful evaluation and selection of external information are essential. The FDA Bayesian guidance [23] outlines several key considerations when assessing the suitability of external data, which are summarized in Table 3. These considerations include data quality and reliability; prespecification of the statistical methods used to incorporate external information (e.g., prior construction); and the relevance of the data based on similarity in estimand attributes, such as eligibility criteria, endpoints, and the handling of intercurrent events. Additional considerations include differences in outcome measurement, the recency of the data, and any important changes over time, such as shifts in the standard of care. With respect to study design, randomized controlled trials are generally preferred over non-randomized studies. Patient-level data are also preferred to aggregate data because they permit a more comprehensive assessment of the relevance and comparability of the external information.

**Table 3:** Criteria for Identifying Available External Information.

| Criteria | Glaze trial | Lavender trial |
| --- | --- | --- |
| **Data quality and reliability** | ✓ | ✓ |
| **Pre-specification** | ✓ | ✓ |
| **Relevance** |  |  |
| – Similarity in estimand attributes | ✓ | ✓ |
| – Any differences in measurement or assessment (e.g., the endpoint) | ✓ | ✓ |
| – Recency of data | ✓ | ✓ |
| – Any potentially important changes | ✓ | ✓ |
| **Study design:** randomized controlled comparisons | ✓ | ✓ |
| **Data availability:** patient-level data | ✗ | ✗ |

We evaluated the external datasets against these criteria and found that both satisfied most of the considerations outlined in the FDA guidance, as indicated in Table 3. Both datasets were obtained from randomized controlled trials, a preferred study design that supports data quality and reliability. In addition, the current study aligns closely with the external studies in key estimand attributes (Table 1), including the target population, efficacy endpoints, and strategies for handling intercurrent events. The external studies are also relatively recent, and no major changes in the standard of care have occurred since their completion. One limitation is that only aggregate data, rather than patient-level data, are available. Nevertheless, the external datasets satisfy most of the relevant criteria and are therefore considered appropriate sources for Bayesian borrowing in the proposed trial.

#### Success criteria

Let Δ = *θ_a_ − θ_p_*, where *θ_a_* and *θ_p_* denote the population mean for an active treatment arm and the placebo arm, respectively, for a given efficacy endpoint. An active treatment arm is declared superior to placebo if the posterior probability that Δ *<* 0 exceeds 0.95 at the final analysis:

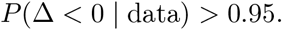

Because lower values of both efficacy endpoints correspond to improved clinical outcomes, a negative value of Δ indicates a favorable treatment effect for the active treatment arm.

#### Interim analysis and futility criteria

Interim futility monitoring is based on the predictive probability of success at the interim analysis, denoted by POS*_I_*. The interim analysis is conducted after 13 participants per treatment arm have completed the study. The predictive probability of success is calculated using the posterior distribution of the treatment effect, *p*(Δ *|* data), while accounting for the remaining participants to be enrolled and followed through the end of the trial:

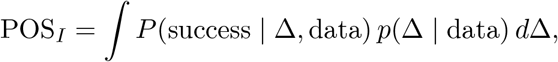

where *P* (success *|* Δ, data) denotes the conditional probability that the prespecified success criterion will be met at the final analysis, given the current interim data. Thus, POS*_I_* represents the posterior predictive probability of ultimately meeting the prespecified success criterion.

An active treatment arm will be discontinued for futility if POS*_I_ < γ* for both efficacy endpoints (RSBQ and CGI-I). Otherwise, enrollment will continue until the planned final sample size of 25 participants per treatment arm is reached.

## 4 Simulation Study

Using the proposed adaptive platform trial in Rett syndrome as a case study, candidate Bayesian borrowing methods were evaluated through simulation to compare their frequentist operating characteristics and inform the selection and prespecification of priors for the proposed design.

### Hierarchical Model Specification

First, we describe the hierarchical model [21] and specify the likelihood, priors, and hyperpriors used in the simulation study.

#### The likelihood

For *i* = 1*, . . ., n* = 25,

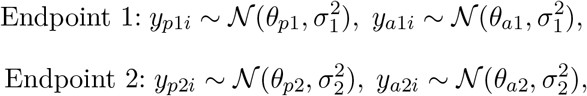

where *y_p_*_1*i*_ and *y_a_*_1*i*_ denote the observed outcomes for subject *i* in the placebo and active treatment arms, respectively, for Endpoint 1, with population means *θ_p_*_1_ and *θ_a_*_1_ and common variance *σ*^2^. The corresponding quantities for Endpoint 2 are defined analogously.

The simulation parameters are set to *θ_p_*_1_ = *−*1.9, *θ_p_*_2_ = 3.7, *σ*_1_ = 9, and *σ*_2_ = 0.75, with values chosen to be broadly consistent with those observed in the external studies (Table 2). The standard deviations, *σ*_1_ and *σ*_2_, were fixed at values consistent with those observed in the external studies and were treated as known in both the simulation and analysis models.

We define *θ_a_*_1_ = *θ_p_*_1_ + Δ_1_ for Endpoint 1 and *θ_a_*_2_ = *θ_p_*_2_ + Δ_2_ for Endpoint 2, where Δ_1_ and Δ_2_ represent the treatment effects for the two endpoints. The values of Δ_1_ and Δ_2_ are specified in the simulation scenarios described below.

#### Prior distributions

We use *N* (0, 100^2^) as a non-informative prior. Although this prior is technically a diffuse (or vague) normal prior, we refer to it as a non-informative prior throughout the paper for simplicity. Non-informative priors are assigned to the active-arm mean parameters, whereas informative priors are specified only for the placebo-arm mean parameters *θ_p_*_1_ and *θ_p_*_2_.

We consider five prior specifications for the control arm: four informative priors representing different degrees of borrowing from the external data summarized in Table 2, and one non-informative prior used as a reference.

1. **MAP**: a MAP prior constructed using the external control data summarized in Table 2.
2. **Mix 20**: an RMAP prior combining the MAP prior with a robust prior component, assigning a weight of 0.2 to the robust component.
3. **Mix 50**: an RMAP prior combining the MAP prior with a robust prior component, assigning a weight of 0.5 to the robust component.
4. **SAM**: a SAM prior with a robust prior component that dynamically adjusts the degree of borrowing based on the agreement between the concurrent placebo data and the external information.
5. **NI**: a non-informative prior, *N* (0, 100^2^).

For the primary analysis, the informative component of all mixture priors is constructed using the same MAP prior. Consequently, MAP, RMAP, and SAM priors differ only in how borrowing from the external information is moderated, allowing a direct comparison of these MAP-based borrowing strategies.

For the robust component, we first consider the non-informative prior, consistent with the original formulations of the RMAP and SAM priors. In addition, we evaluate a skeptical prior as the robust component of the mixture prior in a sensitivity analysis:

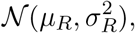

where *µ_R_* is centered at the value corresponding to no symptom improvement, namely *µ_R_* = 0 for Endpoint 1 (RSBQ; ; no change from baseline) and *µ_R_* = 4 for Endpoint 2 (CGI-I; no change at follow-up). The standard deviation *σ_R_* is chosen to be sufficiently small so that large treatment effects are considered unlikely. Specifically, we set *σ_R_* = 4 for Endpoint 1 and *σ_R_* = 1.5 for Endpoint 2, resulting in skeptical priors concentrated around *µ_R_*.

#### Hyperprior distribution

The hyperpriors of *µ* and *τ* are defined as follows:

- 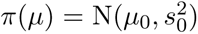, where *µ*_0_ value is no change for each endpoint, while *s*_0_ is chosen to be at least twice the estimated standard deviation [18] from the previous studies.
  1. Endpoint 1: *µ*_0_ = 0 and *s*_0_ = 20.
  2. Endpoint 2: *µ*_0_ = 4 and *s*_0_ = 2.
- *π*(*τ*) = Half-Normal(*σ_τ_*)

Four choices of *σ_τ_* are examined in our simulation study, parameterized by the ratio *F* = *σ/σ_τ_* (with a fixed *σ*). The range of *F* is selected based on rough guidance from the prior maximum sample size, defined as the ratio of within-trial to between-trial variance, *n^∗^* = *σ*^2^*/τ* ^2^, which under fixed values of *σ* and *τ*, can be used as a scale for quantifying between-trial heterogeneity [12].

### Simulation Scenarios

#### Treatment effects

Let Δ*_i_* denote the treatment-effect for Endpoint *i*, *i* = 1, 2. We consider three treatment-effect scenarios, including a null scenario with no treatment effect, as summarized in Table 4. The treatment effects combinations (Δ_1_, Δ_2_) = (*−*3.5*, −*0.25) and (Δ_1_, Δ_2_) = (*−*5.0*, −*0.35) represent moderate and large treatment effects, respectively. The moderate treatment effect is broadly comparable to that observed in previous studies.

**Table 4:** Treatment Effects.

| Endpoint | $\Delta$ | Scenario 1 | Scenario 2 | Scenario 3 |
| --- | --- | --- | --- | --- |
| Endpoint 1 | $\Delta_1$ | 0 | -3.5 | -5.0 |
| Endpoint 2 | $\Delta_2$ | 0 | -0.25 | -0.35 |

**Table 5:** Drift for Prior–Data Conflict.

| Endpoint | $\hat{\mu} + 2d$ | $\hat{\mu} + d$ | $\hat{\mu}$ | $\hat{\mu} - d$ | $\hat{\mu} - 2d$ |
| --- | --- | --- | --- | --- | --- |
| Endpoint 1 | $\hat{\mu} + 4$ | $\hat{\mu} + 2$ | $\hat{\mu}$ | $\hat{\mu} - 2$ | $\hat{\mu} - 4$ |
| Endpoint 2 | $\hat{\mu} + 0.5$ | $\hat{\mu} + 0.25$ | $\hat{\mu}$ | $\hat{\mu} - 0.25$ | $\hat{\mu} - 0.5$ |

**Table 6:** Clinically Significant Difference (CSD).

| Endpoint | CSD 1 | CSD 2 | CSD 3 |
| --- | --- | --- | --- |
| Endpoint 1 | -2.0 | -3.5 | -6.0 |
| Endpoint 2 | -0.10 | -0.25 | -0.50 |

#### Prior–data conflict

We introduce a drift parameter, *d*, to represent the degree of prior–data conflict. The values *d* = 2 and *d* = 0.25 are used for Endpoint 1 and Endpoint 2, respectively. The drift is defined relative to *µ*^, the estimated mean of the MAP prior constructed from the external studies, so that *µ*^ corresponds to the absence of prior–data conflict. Positive and negative values of *d* represent departures from the mean of external data in opposite directions, with larger absolute values of *d* indicating greater disagreement between the current and external data.

#### Between-study heterogeneity (or similarity)

The prior for the between-study standard deviation, *τ*, is parameterized in terms of *F*, a parameter that reflects the degree of between-study similarity. We consider four values of *F* (2, 3, 4, and 8) to evaluate sensitivity to the assumed degree of between-study similarity. For a fixed within-study standard deviation, *σ*:

- Larger values of *F* imply smaller values of *τ*, indicating greater similarity among studies and therefore stronger borrowing.
- Smaller values of *F* imply larger values of *τ*, indicating greater between-study heterogeneity and therefore weaker borrowing.

#### Clinically significant difference

The SAM prior requires specification of a clinically significant difference (CSD) *a priori*, which is used to define the alternative hypothesis in the adaptive borrowing procedure. We examine sensitivity to three prespecified CSD values corresponding to small (CSD 1), moderate (CSD 2), and large (CSD 3) treatment effects.

### Simulation design summary and evaluation metrics

In summary, we compare five prior specifications (MAP, Mix 20, Mix 50, SAM, and NI) across 180 simulation scenarios, corresponding to all combinations of:

- 3 treatment-effect scenarios,
- 5 levels of prior–data conflict,
- 4 values of *F* representing between-study heterogeneity, and
- 3 CSD values for the SAM prior.

For each simulation scenario, 1,000 independent trial replicates were generated to estimate the frequentist operating characteristics. Primary evaluation metrics include bias, relative mean squared error (rMSE), and the probability of success at the final analysis, which corresponds to the Type I error rate under the null hypothesis and statistical power under the alternative hypothesis. The rMSE is defined relative to the NI prior as the relative difference between the MSE obtained using each informative prior and that obtained using the NI prior, 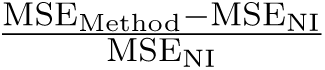. Additional evaluation metrics include the probability of early discontinuation of an active treatment arm at the interim analysis, the weight assigned to the robust component of the mixture prior, and the effective sample size (ESS).

### Simulation Results

The choice of the CSD had only a modest impact on the frequentist operating characteristics (see the illustrative example in Figure S1), although it substantially affected the degree of borrowing under the SAM prior, as reflected by the ESS (Figure S2). Therefore, all subsequent results are presented using a fixed CSD of *−*3.5 for Endpoint 1 and *−*0.25 for Endpoint 2 (CSD 2).

We first evaluated sensitivity to the between-study similarity parameter *F*, which determines the prior distribution of the between-study standard deviation *τ*, and selected *F* = 4 because it provided a reasonable balance between borrowing efficiency and robustness across the simulation scenarios.

Unless otherwise stated, we present results for Endpoint 1 because Endpoint 2 exhibited very similar trends (see the Supplementary Material, Figures S3–S8). Results shown in the main text use a non-informative prior as the robust component of the mixture prior, whereas the corresponding sensitivity analyses using a skeptical prior are presented in the Supplementary Material (Figures S9– S14).

Throughout the figures, the horizontal axis represents the degree of prior–data conflict (drift). The center, *µ*^, corresponds to no prior–data conflict, and increasing distance from *µ*^ in either direction indicates greater disagreement between the external and current data.

#### Bias and relative mean squared error (rMSE)

Bias and rMSE are shown in Figures 1 and 2. As expected, NI exhibited essentially no bias across all scenarios. In contrast, informative borrowing introduced bias under prior–data conflict, with the magnitude of bias increasing as the drift moved farther from *µ*^. The MAP prior showed the largest bias under substantial conflict, whereas the robust mixture priors mitigated this effect. Among the robust approaches, Mix 20 and Mix 50 produced intermediate bias, while the SAM prior generally remained closer to NI than either Mix 20 or Mix 50.

**Figure 1:**
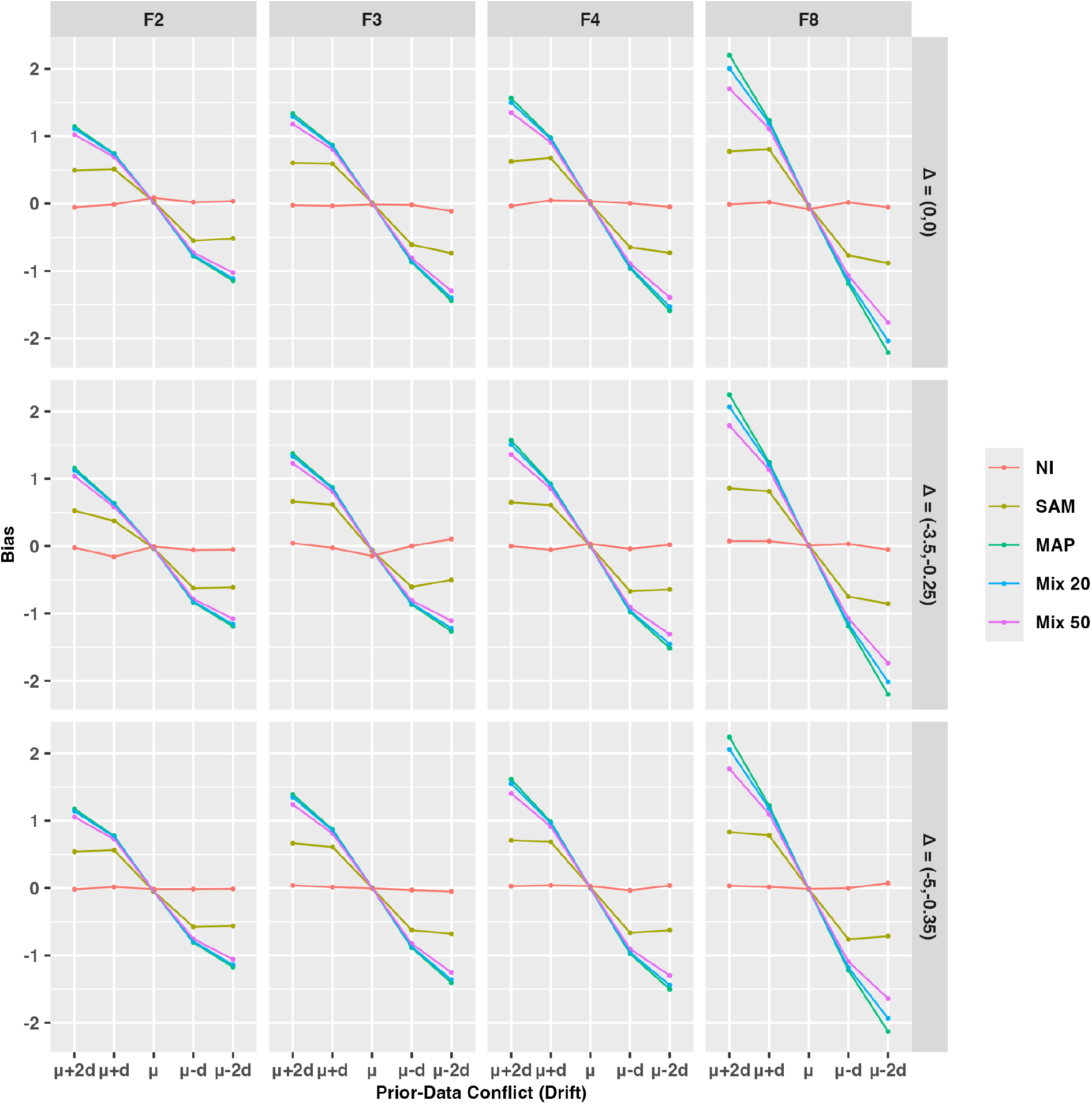
Bias for Endpoint 1 as a function of prior–data conflict (drift) and the heterogeneity prior. Results are shown for the primary analysis using a non-informative prior as the robust component of the mixture prior. Columns correspond to the values of *F* used to specify the prior distribution for the between-study heterogeneity parameter *τ*, and rows correspond to the three treatment-effect scenarios.

**Figure 2:**
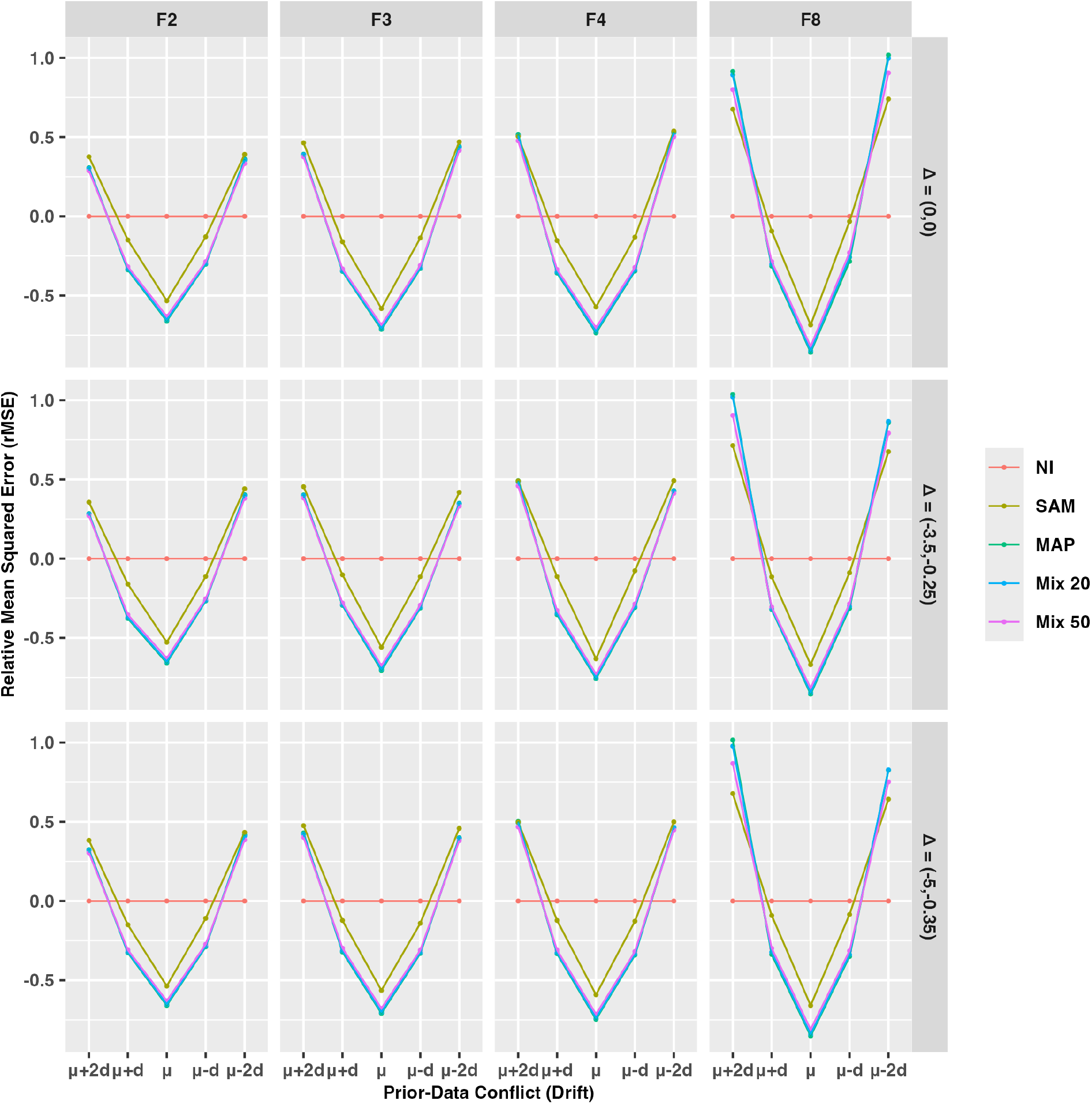
Relative mean squared error (rMSE) for Endpoint 1 as a function of prior–data conflict (drift) and the heterogeneity prior. Results are shown for the primary analysis using a non-informative prior as the robust component of the mixture prior. The relative MSE is calculated with respect to the non-informative prior (NI), such that values below zero indicate improved estimation efficiency compared with NI. Columns correspond to the values of *F* used to specify the prior distribution for the between-study heterogeneity parameter *τ*, and rows correspond to the three treatment-effect scenarios.

The rMSE demonstrates the expected bias–variance tradeoff. Relative to NI (rMSE = 0), all borrowing approaches reduced MSE when prior–data conflict was absent or modest, reflecting variance reduction through borrowing. As prior–data conflict increased, this benefit gradually diminished and eventually reversed as increasing bias outweighed the reduction in variance. The MAP prior showed the greatest deterioration under severe conflict, whereas the robust mixture priors were more resistant. Under the primary analysis using a non-informative robust component, differences among MAP, Mix 20, and Mix 50 were relatively modest. In the sensitivity analysis using a skeptical robust component, the separation between these methods became more pronounced, with progressively greater robustness observed from MAP to Mix 20, Mix 50, and SAM (see the Supplementary Material, Figures S9 and S10).

#### Probability of early discontinuation of an active treatment arm, Type I error, and power

Figures 3 and 4 summarize the frequentist operating characteristics of the adaptive design. Under the null scenario (Δ = (0, 0)), the probability of early discontinuation of an active treatment arm at the interim analysis was generally high, except when substantial drift favored the active treatment and increased the apparent treatment difference between the active and placebo arms. Early discontinuation of futile treatment arms helped maintain control of the Type I error rate across most simulation scenarios.

**Figure 3:**
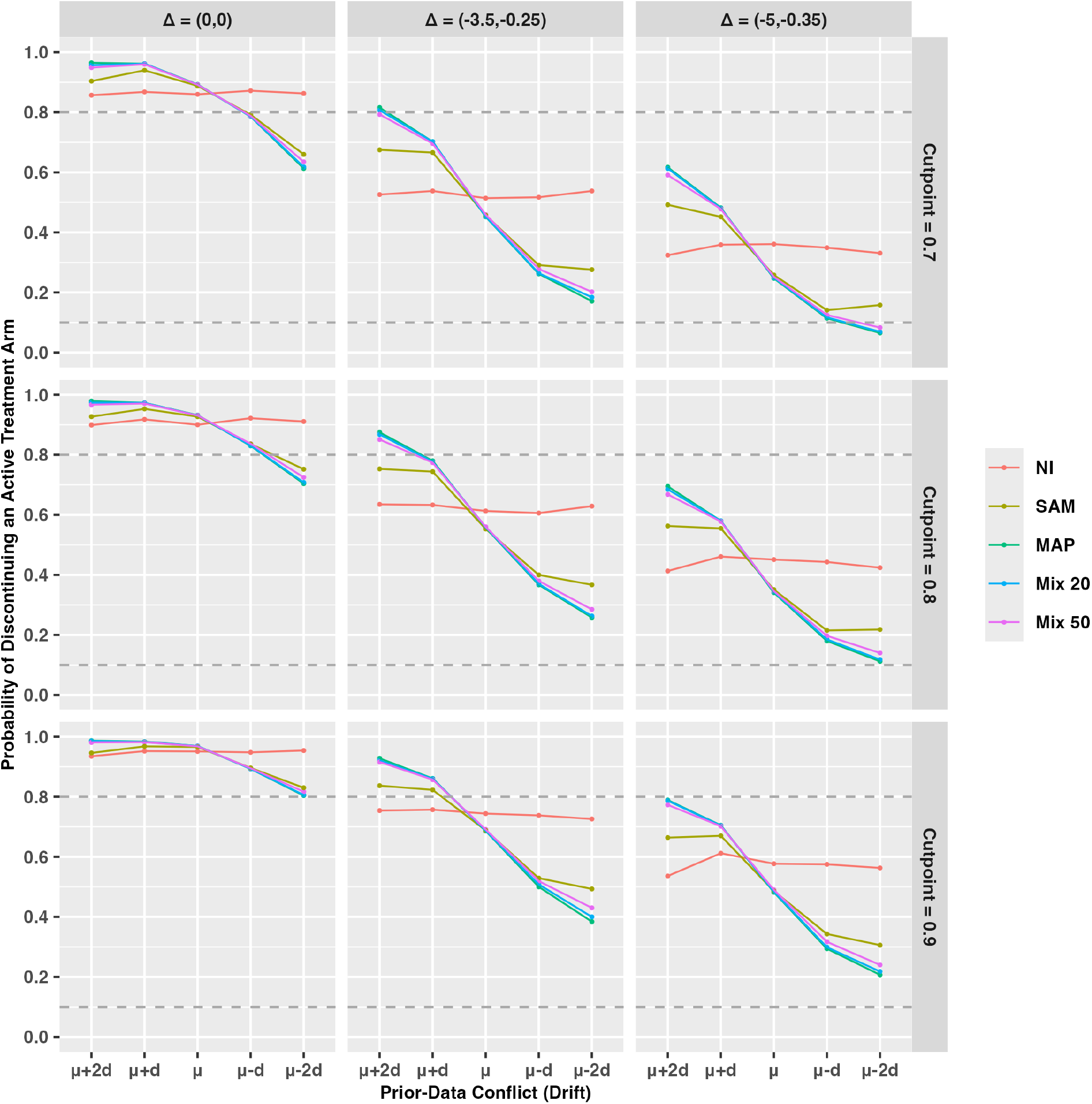
Probability of early discontinuation of an active treatment arm at the interim analysis for Endpoint 1 as a function of prior–data conflict (drift) and treatment-effect scenario. Results are shown for the primary analysis using a non-informative prior as the robust component of the mixture prior. Columns correspond to the three treatment-effect scenarios, and rows correspond to the futility cutoff values (*γ* = 0.7, 0.8, and 0.9) used at the interim analysis.

**Figure 4:**
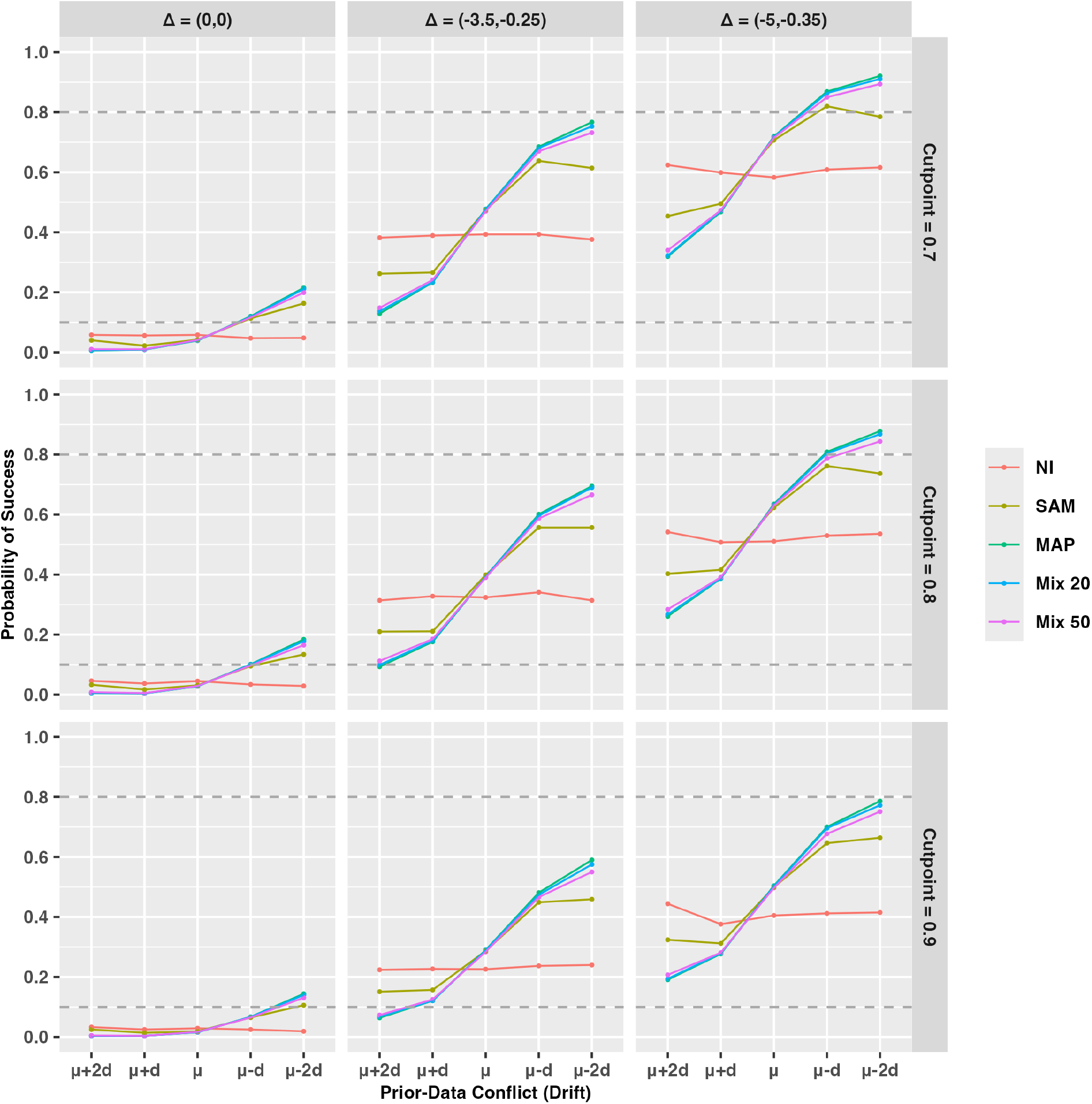
Probability of success at the final analysis for Endpoint 1 as a function of prior–data conflict (drift) and treatment-effect scenario. Results are shown for the primary analysis using a non-informative prior as the robust component of the mixture prior. Under the null scenario (i.e., Δ = (0, 0)), the probability of success represents the Type I error rate, whereas under the alternative scenarios it represents statistical power. The horizontal dashed lines indicate the nominal Type I error rate (0.10) and the target power (0.80). Columns correspond to the three treatment-effect scenarios, and rows correspond to the futility cutoff values (*γ* = 0.7, 0.8, and 0.9) used at the interim analysis.

Under the alternative scenarios, the probability of success depended strongly on both the magnitude and direction of prior–data conflict (Figure 4). When the external information favored the active treatment, borrowing increased the probability of success relative to NI, whereas conflict in the opposite direction reduced the probability of success. The differences among the borrowing methods became more pronounced as prior–data conflict increased. The MAP prior generally exhibited the largest departure from NI, followed by Mix 20 and Mix 50, while the SAM prior generally remained closest to NI across the range of prior–data conflict considered, reflecting its adaptive reduction in borrowing under prior–data conflict.

Similar qualitative patterns were observed in the sensitivity analysis using a skeptical prior as the robust component of the mixture prior. However, the differences among the borrowing methods became more pronounced, particularly between the MAP prior and the RMAP priors (Mix 20 and Mix 50), whereas the SAM prior remained closest to NI across the range of prior–data conflict considered (see the Supplementary Material, Figures S11 and S12).

#### Weight assigned to the robust component

Figure 5 presents the average weight assigned to the robust component of the mixture prior at the interim and final analyses across the simulated trials. For the RMAP priors, these weights are fixed by design and are shown for reference. In contrast, the SAM prior estimates the weight adaptively from the current data. The SAM prior assigns the smallest weight to the robust component when little or no prior–data conflict is present, thereby allowing the greatest degree of borrowing from the informative prior. As prior–data conflict increases, the weight assigned to the robust component also increases, reducing the influence of the external information. This adaptive behavior becomes more pronounced at the final analysis because additional current data provide stronger evidence regarding the agreement between the external and current data.

**Figure 5:**
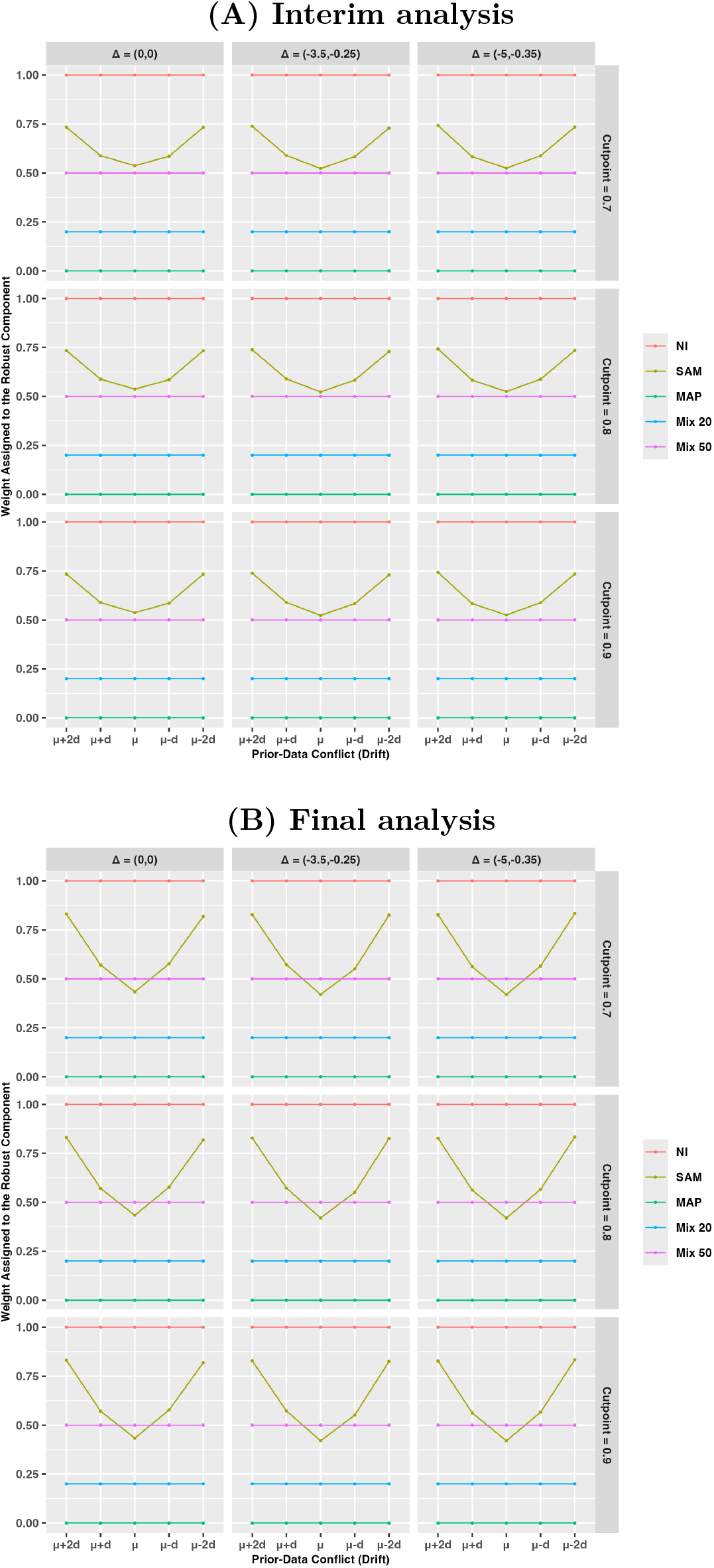
Weight assigned to the robust component of the mixture prior for Endpoint 1. Results are shown for the primary analysis using a non-informative prior as the robust component of the mixture prior. Panel (A) shows the weight at the interim analysis, and Panel (B) shows the weight at the final analysis. For the RMAP priors, the weights are fixed by design, whereas for the SAM prior, the weight is adaptively estimated from the observed data. Columns correspond to the three treatment-effect scenarios, and rows correspond to the futility cutoff values (*γ* = 0.7, 0.8, and 0.9) used at the interim analysis.

#### Effective sample size

Figure 6 shows the average ESS contributed by the external information across the simulated trials using CSD 2. Consistent with its adaptive weighting mechanism, the ESS for the SAM prior is highest when prior–data conflict is absent and decreases as conflict increases. In contrast, the ESS remains constant for the MAP and RMAP priors because their borrowing weights are fixed. Under CSD 2, the maximum ESS contributed by the SAM prior at the final analysis is approximately 15 for Endpoint 1, substantially smaller than the external sample size of 109. Figure S2 demonstrates that increasing the CSD results in greater adaptive borrowing and, consequently, a larger ESS under the SAM prior, approaching the ESS of the MAP prior under complete agreement between the external and current data. These results illustrate that the SAM prior adaptively limits borrowing while preserving meaningful efficiency gains when the external and current data are compatible.

**Figure 6:**
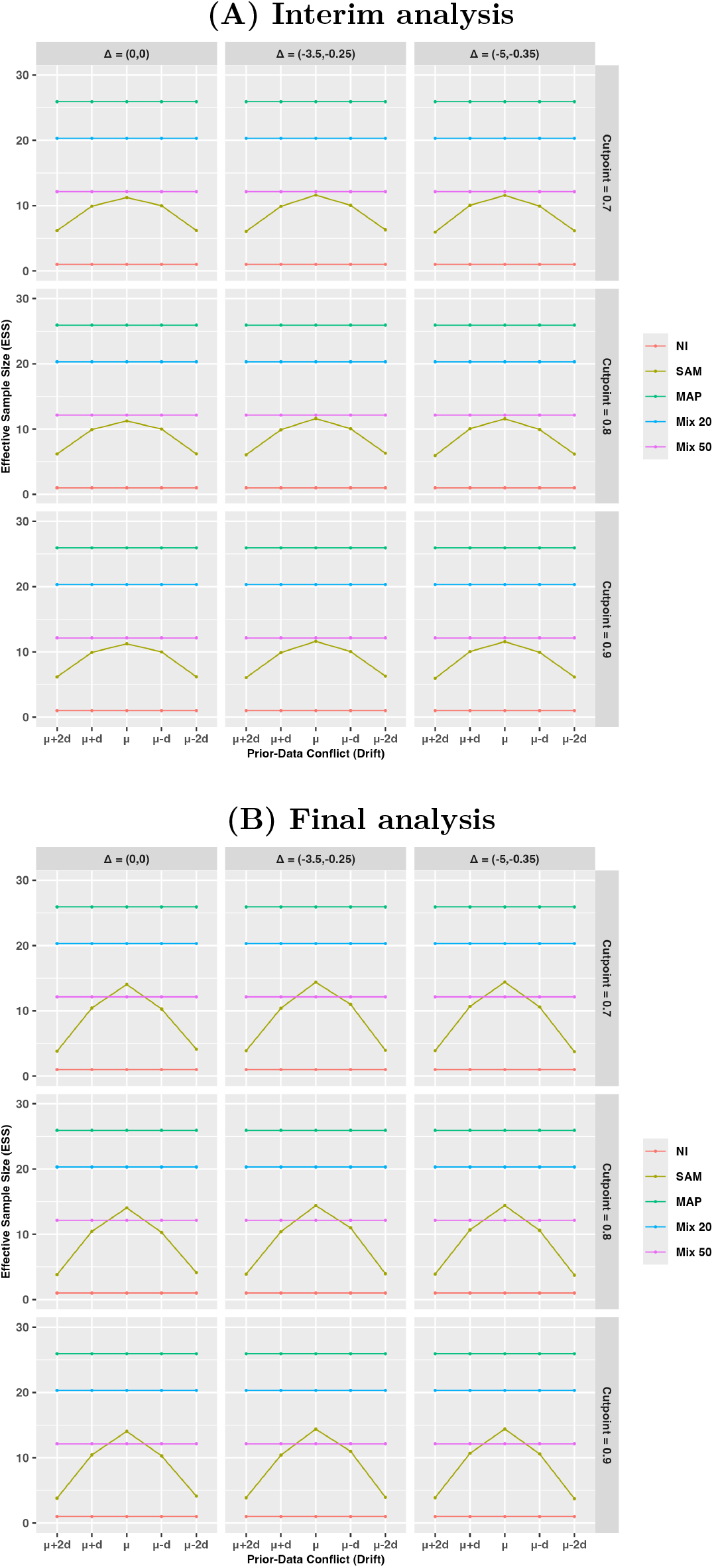
Effective sample size (ESS) contributed by the external information for Endpoint 1. Results are shown for the primary analysis using a non-informative prior as the robust component of the mixture prior. Panel (A) shows the ESS at the interim analysis, and Panel (B) shows the ESS at the final analysis. Columns correspond to the three treatment-effect scenarios, and rows correspond to the futility cutoff values (*γ* = 0.7, 0.8, and 0.9) used at the interim analysis.

## 5 Discussion

Bayesian borrowing of external information has the potential to improve the efficiency of clinical trials by reducing the number of concurrently enrolled control participants [20], an important consideration in rare disease research where patient populations are limited. However, borrowing external information also introduces potential risks, including increased bias and inflation of the Type I error rate, when the current trial differs from the external studies [3, 24]. Consequently, a central challenge in Bayesian trial design is balancing statistical efficiency with robustness when incorporating external information [19]. Motivated by these challenges and recent regulatory emphasis on rigorous evaluation of external information [23], we compared several MAP-based borrowing strategies that differ in how they moderate borrowing from external information and evaluated their frequentist operating characteristics using a simulation study motivated by an adaptive platform trial in Rett syndrome.

The simulation results demonstrate the expected bias–variance tradeoff associated with Bayesian borrowing. Under little or no prior–data conflict, informative priors reduced variance and increased the probability of success relative to a non-informative prior. As disagreement between the external and current data increased, informative borrowing became increasingly susceptible to bias, illustrating the importance of accounting for potential non-exchangeability [13, 19, 24]. The MAP prior achieved the greatest efficiency when the external and current data were compatible but also exhibited the largest bias and the greatest departures from the non-informative analysis under substantial prior–data conflict [26]. The RMAP priors mitigated these effects by combining the informative MAP prior with a robust component using pre-specified mixture weights [19], while the SAM prior further improved robustness by adaptively adjusting the degree of borrowing according to the observed agreement between the concurrent placebo data and the external information [29]. Across the simulation scenarios considered, the SAM prior remained closest to the non-informative analysis while retaining meaningful efficiency gains when the external and current data were compatible.

Although the RMAP prior is sometimes described as adaptive because the influence of the informative component is reduced through Bayesian updating when it conflicts with the current data, the borrowing mechanism itself is fixed once the mixture weight has been specified *a priori*. Consequently, the RMAP prior is not a dynamic borrowing approach, as noted by Yang et al. [29]. Instead, its robustness arises from posterior updating, which naturally discounts the informative component when it is inconsistent with the current data [19]. In contrast, the SAM prior explicitly estimates the mixture weight from the observed data, allowing the degree of borrowing to adapt according to the compatibility between the external and current information. Thus, the RMAP prior requires investigators to specify how much confidence to place in the external information before observing the current data, whereas the SAM prior allows the observed data to determine the extent to which the external information should be borrowed.

The simulation results further illustrate the consequences of prior–data conflict. When the direction of the conflict favored the active treatment, borrowing increased the probability of success relative to the non-informative analysis. Conversely, when the external information suggested better clinical outcomes for the placebo arm (e.g., larger placebo effects than observed in the current trial), borrowing reduced the probability of success. These findings emphasize that both the magnitude and direction of prior–data conflict influence trial operating characteristics [24] and reinforce the importance of evaluating a broad range of conflict scenarios during trial planning [23]. The increasing availability of user-friendly software further facilitates the practical implementation of Bayesian borrowing methods. For example, the RBesT package provides comprehensive tools for constructing MAP and RMAP priors [27], while the SAMprior package implements the SAM prior framework [28]. These software tools are expected to facilitate broader adoption of Bayesian methods incorporating external information in future clinical trials.

The sensitivity analysis demonstrated that replacing the non-informative prior with a skeptical prior as the robust component of the mixture prior did not alter the qualitative conclusions. Rather, the differences among the borrowing methods became more pronounced. Because the skeptical prior is more conservative than the diffuse non-informative prior, it more strongly penalizes borrowing when disagreement between the external and current data arises. Consequently, the separation between the MAP prior and the RMAP priors increased, whereas the adaptive behavior of the SAM prior remained largely unchanged. These findings indicate that the main conclusions are robust to the specification of the robust component.

Our findings reinforce the well-established role of the heterogeneity prior in MAP-based borrowing [12, 26]. Although the prior for the overall mean is typically weakly influential, the prior placed on the between-study heterogeneity parameter, *τ*, substantially affects the degree of borrowing by reflecting the assumed similarity among studies. Consistent with the Bayesian borrowing literature, our simulation study illustrates that the choice of the heterogeneity prior can meaningfully influence operating characteristics [26] and therefore should be carefully justified during trial planning. In contrast, the CSD used by the SAM prior primarily influenced the degree of adaptive borrowing, as reflected by the ESS. Although larger CSD values resulted in greater borrowing, the resulting changes in the frequentist operating characteristics were modest across the simulation scenarios considered.

Several limitations should be acknowledged. First, the simulation study was based on aggregated external data. Patient-level external data would permit more flexible borrowing strategies and a more comprehensive assessment of exchangeability between the external and current populations [3, 23]. Second, although this work focused on MAP-based borrowing strategies, other Bayesian borrowing approaches, such as power priors [7], commensurate priors [6], calibrated power priors [15], elastic priors [8], and latent exchangeability priors [1], represent alternative frameworks that warrant future investigation. Finally, the simulation study focused on normally distributed continuous endpoints within a single adaptive platform trial setting. Although these settings were motivated by the Rett syndrome case study, further evaluation is needed to determine the generalizability of the findings to other endpoint types and adaptive trial designs.

Future work will extend these methods to patient-level external data. One of the main strengths of the meta-analytic approach is that it can accommodate both patient-level and aggregate external data [20]. Another promising direction is the joint modeling of patient-level external and concurrent trial data using bias parameters to account for potential systematic differences between the data sources. Such approaches may allow borrowing of information on precision while accommodating differences in treatment effects when there is reasonable suspicion of bias [23]. When patient-level external data are available, propensity score methods [17, 22] and regression-based covariate adjustment can be used to improve exchangeability between external and concurrent control populations. More recently, methods that combine propensity score methods with Bayesian borrowing approaches, including power priors and MAP priors, have been proposed to better account for between-trial heterogeneity [9, 10, 16, 25, 30]. Evaluating the impact of propensity score methods on the frequentist operating characteristics of Bayesian borrowing approaches represents an important direction for future research. Extending the proposed framework to additional end-point types and more complex adaptive trial designs, including multiple interim analyses, treatment selection, and response-adaptive randomization, also represents an important direction for future methodological research.

The increasing availability of external data, together with evolving regulatory environment, has created new opportunities for incorporating external information into clinical trial design. The recent FDA draft Bayesian guidance emphasizes careful evaluation of external data sources, pre-specification of Bayesian borrowing methods, and assessment of operating characteristics under plausible prior–data conflict scenarios. The framework presented in this paper aligns with these recommendations by combining systematic evaluation of external information, transparent specification of MAP-based borrowing strategies, and comprehensive simulation studies to quantify the frequentist operating characteristics before trial initiation [23]. Such an approach provides a practical framework for designing robust Bayesian clinical trials that efficiently leverage external information while maintaining appropriate operating characteristics under uncertainty regarding exchangeability.

## Acknowledgements

During the preparation of this manuscript, the authors used ChatGPT (OpenAI) to assist with language editing to improve clarity and refine the manuscript. ChatGPT was not used to conduct any analyses or generate the study results. The use of ChatGPT did not alter the study results or conclusions. The authors take full responsibility for the content of the manuscript.

## Data Availability

The external summary data used to inform the simulation study were obtained from previously published studies cited in this article.

## Ethics Statement

No ethics approval was required for this study, which was based on simulation studies and previously published aggregate data.

## Patient Consent Statement

Not applicable.

## Figures

**Abbreviations used throughout the figures.** NI: non-informative prior, *N* (0, 100^2^); SAM: self-adapting mixture prior with an adaptively determined robust-component weight; MAP: meta-analytic-predictive prior incorporating external control data; Mix 20: RMAP prior with a fixed robust-component weight of 0.20; Mix 50: RMAP prior with a fixed robust-component weight of 0.50.

