## Supplementary Material for "Bayesian Borrowing of External Information in Clinical Trials: A Comparison of MAP, RMAP, and SAM Priors"

This supplement provides additional simulation results supporting the primary analyses presented in the main manuscript. Figure S1 examines the sensitivity of the SAM prior to the choice of clinically significant difference (CSD), while Figure S2 shows the effective sample size (ESS) contributed by the external information for Endpoint 1 under two alternative CSD values for the SAM prior. Figures S3–S8 present additional results for Endpoint 2 under the primary analysis using a non-informative prior as the robust component of the mixture prior, whereas Figures S9–S14 present sensitivity analyses for Endpoint 1 using a skeptical prior as the robust component of the mixture prior.

**Abbreviations used throughout the supplementary figures.**

NI: non-informative prior, $\mathcal{N}(0, 100^2)$

SAM: self-adapting mixture prior with an adaptively determined robust-component weight

MAP: meta-analytic-predictive prior incorporating external control data

Mix 20: RMAP prior with a fixed robust-component weight of 0.20

Mix 50: RMAP prior with a fixed robust-component weight of 0.50

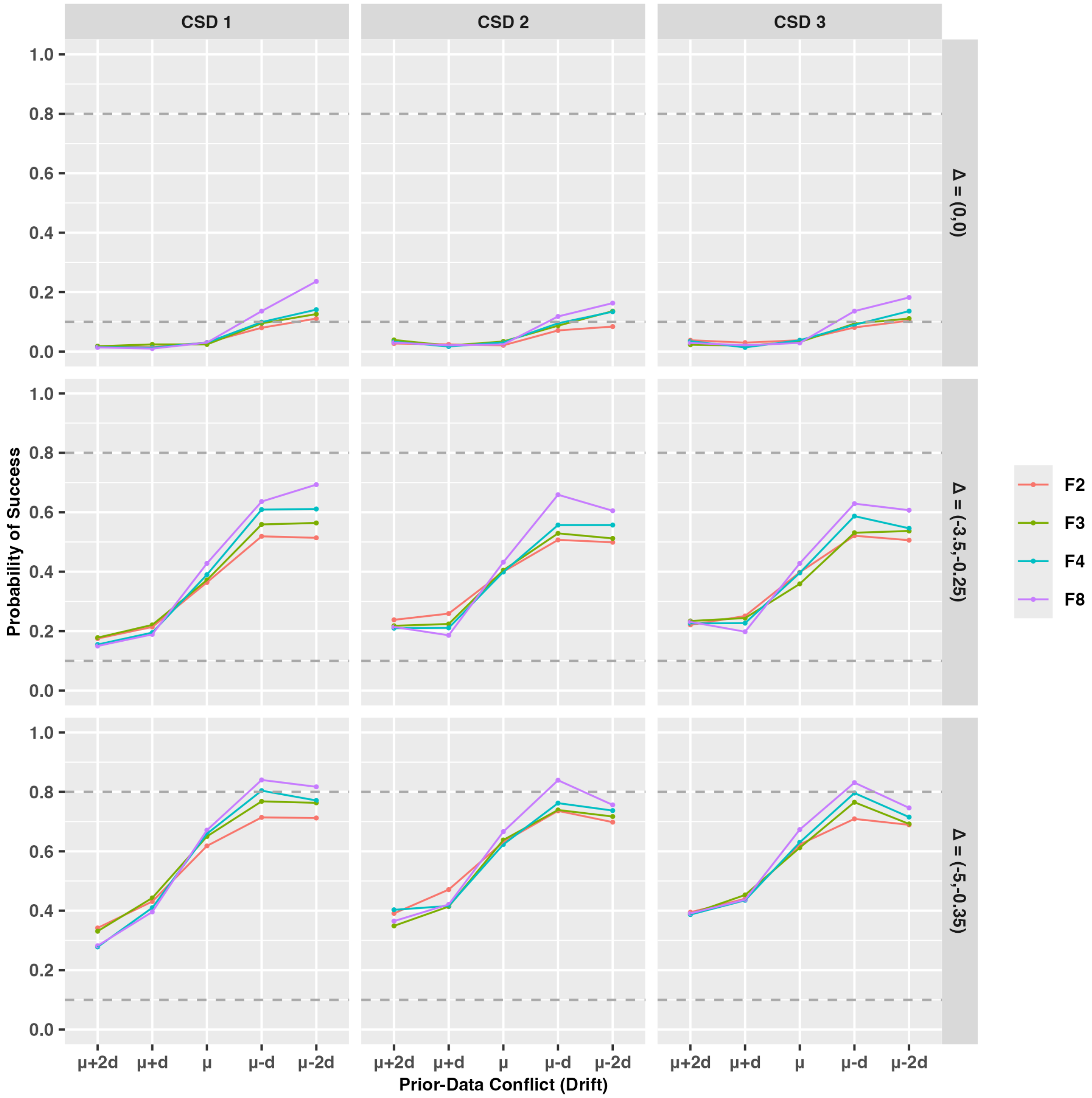


**Figure S1:** Probability of success at the final analysis for Endpoint 1 as a function of the clinically significant difference (CSD) under the SAM prior. Results are shown for the primary analysis using a non-informative prior as the robust component of the mixture prior. Under the null scenario ($\Delta = (0, 0)$), the probability of success corresponds to the Type I error rate, whereas under the alternative scenarios it corresponds to statistical power. Columns correspond to the three CSD values, rows correspond to the treatment-effect scenarios, and colors denote the values of $F$ used to specify the prior distribution for $\tau$. The futility decision at the interim analysis was based on a cutoff value of $\gamma = 0.8$. These results indicate that the operating characteristics were largely insensitive to the choice of CSD across the values of $F$ considered.

**(A) Clinically significant differences (CSD 1):** $(-2.0, -0.10)$

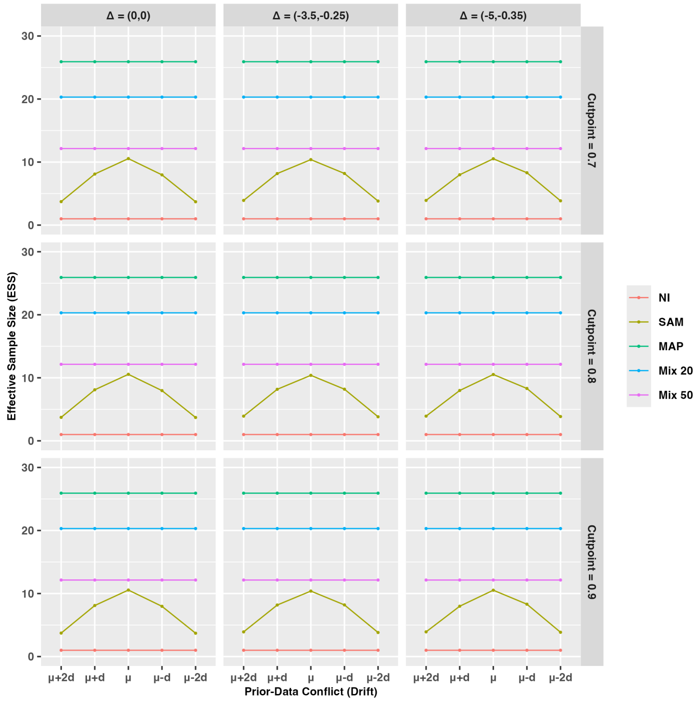


**(B) Clinically significant differences (CSD 3):** $(-6.0, -0.50)$

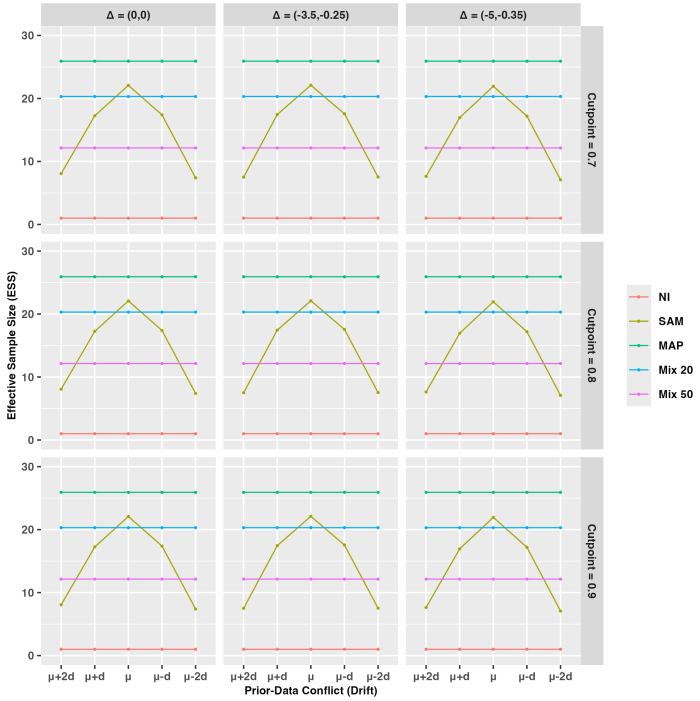


**Figure S2:** Effective sample size (ESS) contributed by the external information for Endpoint 1. Results are shown for the primary analysis using a non-informative prior as the robust component of the mixture prior. Panel (A) shows the ESS using clinically significant differences (CSD) of $(-2.0, -0.10)$ (CSD 1), and Panel (B) shows the ESS using CSD of $(-6.0, -0.50)$ (CSD 3). Columns correspond to the three treatment-effect scenarios, and rows correspond to the futility cutoff values ($\gamma$ = 0.7, 0.8, and 0.9) used at the interim analysis. A larger CSD results in greater adaptive borrowing under the SAM prior, as reflected by a larger ESS.

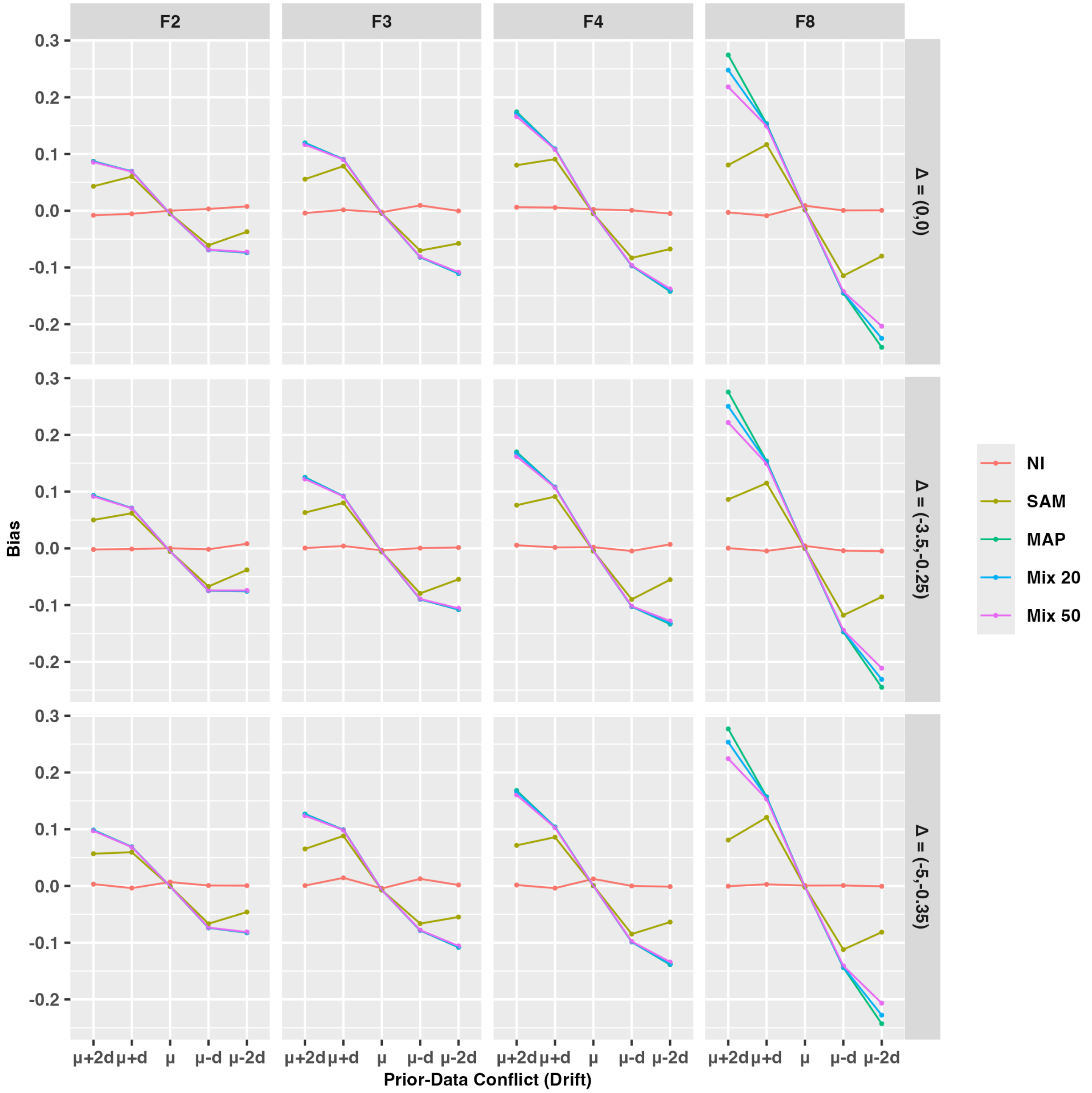


**Figure S3:** Bias for Endpoint 2 as a function of prior–data conflict (drift) and the heterogeneity prior. Results are shown for the primary analysis using a non-informative prior as the robust component of the mixture prior. Columns correspond to the values of $F$ used to specify the prior distribution for the between-study heterogeneity parameter $\tau$, and rows correspond to the three treatment-effect scenarios.

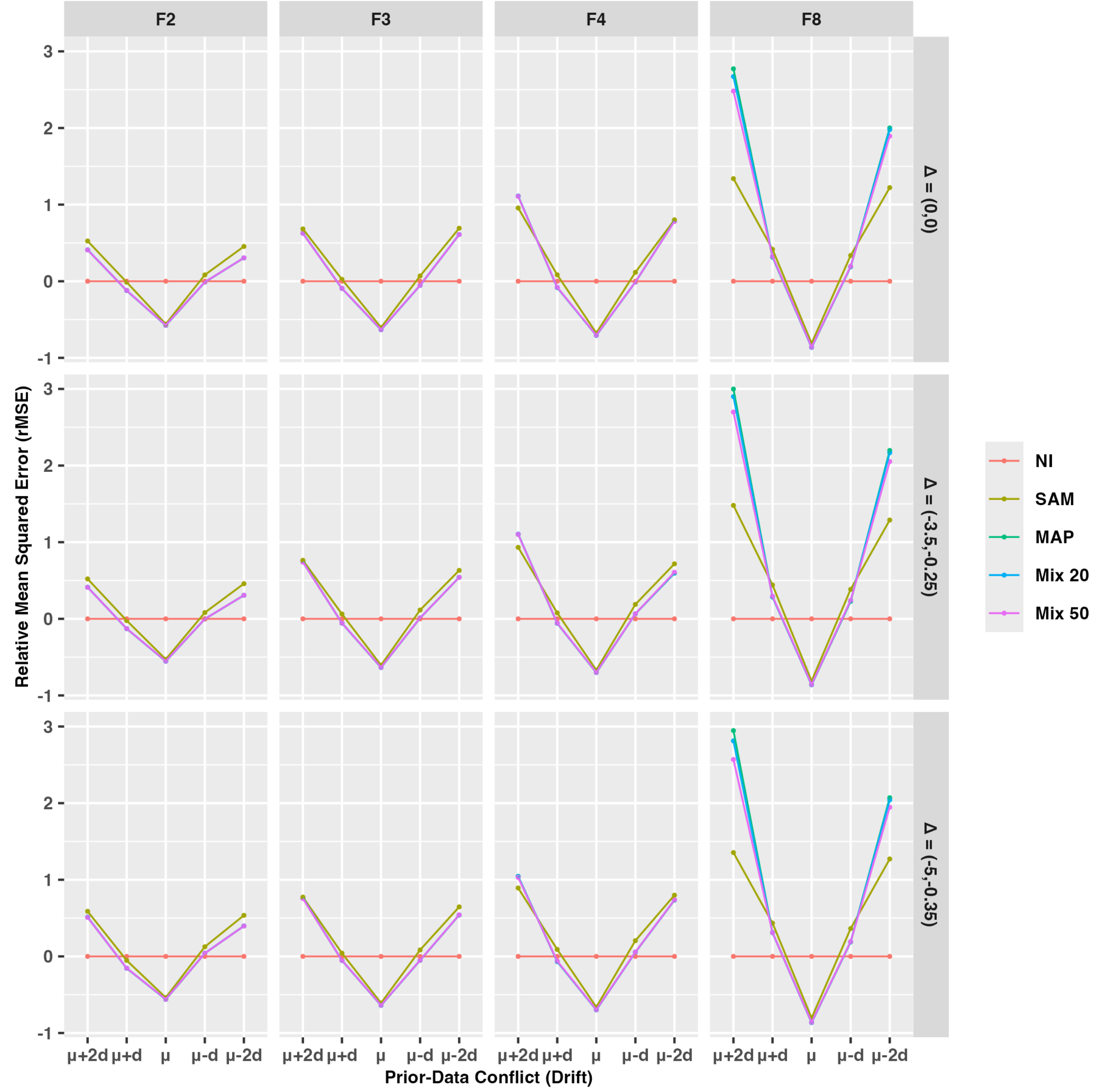


**Figure S4:** Relative mean squared error (rMSE) for Endpoint 2 as a function of prior–data conflict (drift) and treatment-effect scenario. Results are shown for the primary analysis using a non-informative prior as the robust component of the mixture prior. The relative MSE is calculated with respect to the non-informative prior (NI), such that values below zero indicate improved estimation efficiency compared with NI. Columns correspond to the values of $F$ used to specify the prior distribution for the between-study heterogeneity parameter $\tau$, and rows correspond to the three treatment-effect scenarios.

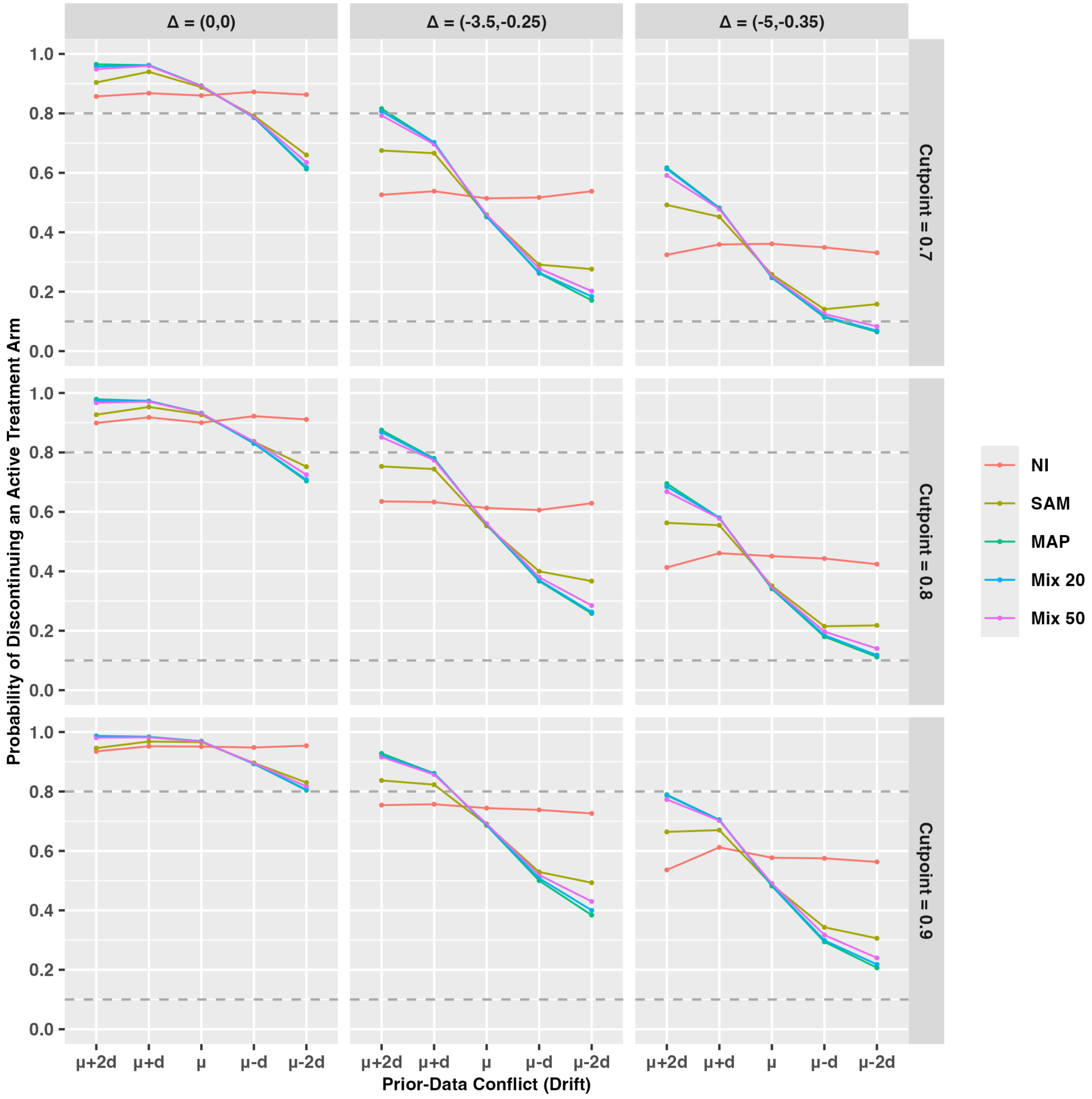


**Figure S5:** Probability of early discontinuation of an active treatment arm at the interim analysis for Endpoint 2 as a function of prior–data conflict (drift) and treatment-effect scenario. Results are shown for the primary analysis using a non-informative prior as the robust component of the mixture prior. Columns correspond to the three treatment-effect scenarios, and rows correspond to the futility cutoff values ($\gamma = 0.7$, 0.8, and 0.9) used at the interim analysis.

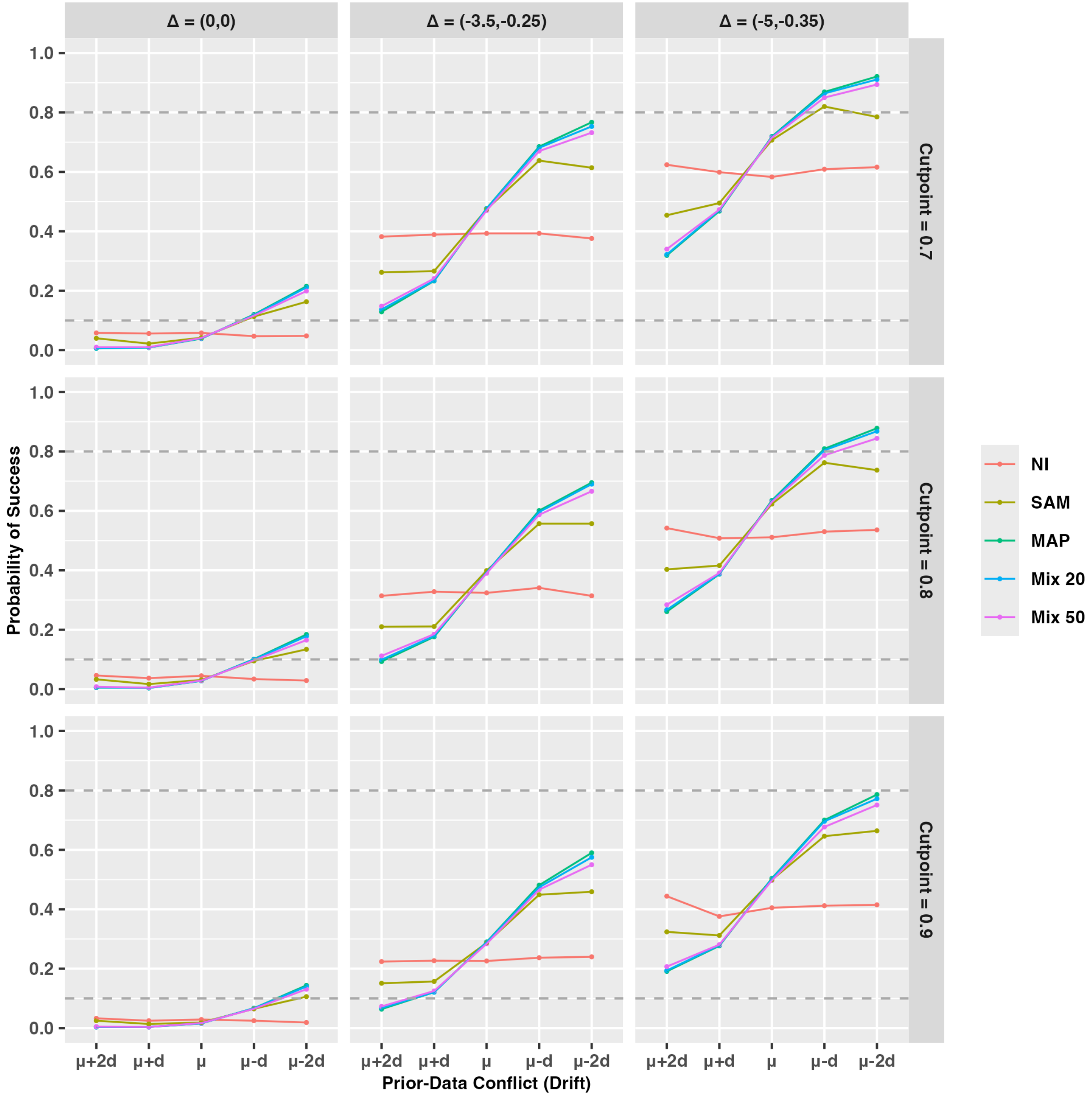


**Figure S6:** Probability of success at the final analysis for Endpoint 2 as a function of prior–data conflict (drift) and treatment-effect scenario. Results are shown for the primary analysis using a non-informative prior as the robust component of the mixture prior. Under the null scenario (i.e., $\Delta = (0, 0)$), the probability of success represents the Type I error rate, whereas under the alternative scenarios it represents statistical power. The horizontal dashed lines indicate the nominal Type I error rate (0.10) and the target power (0.80). Columns correspond to the three treatment-effect scenarios, and rows correspond to the futility cutoff values ($\gamma$ = 0.7, 0.8, and 0.9) used at the interim analysis.

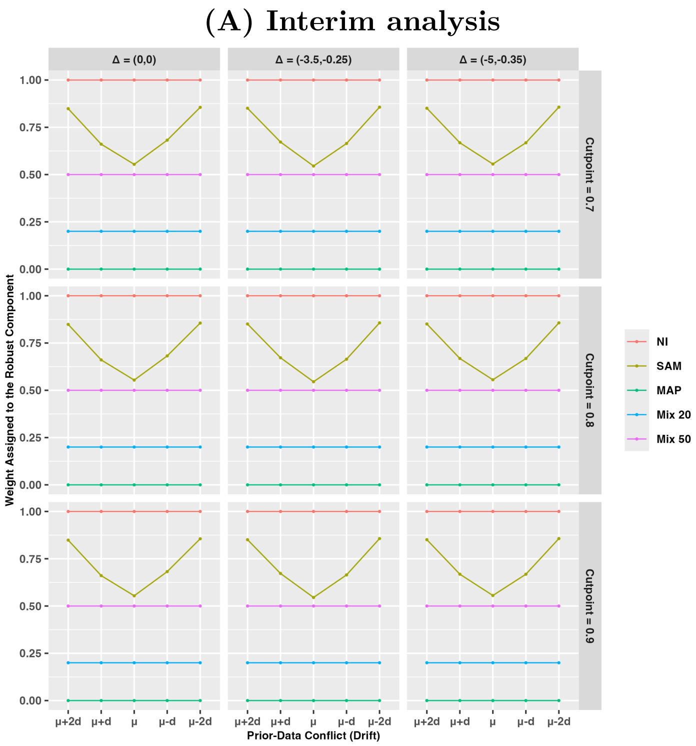


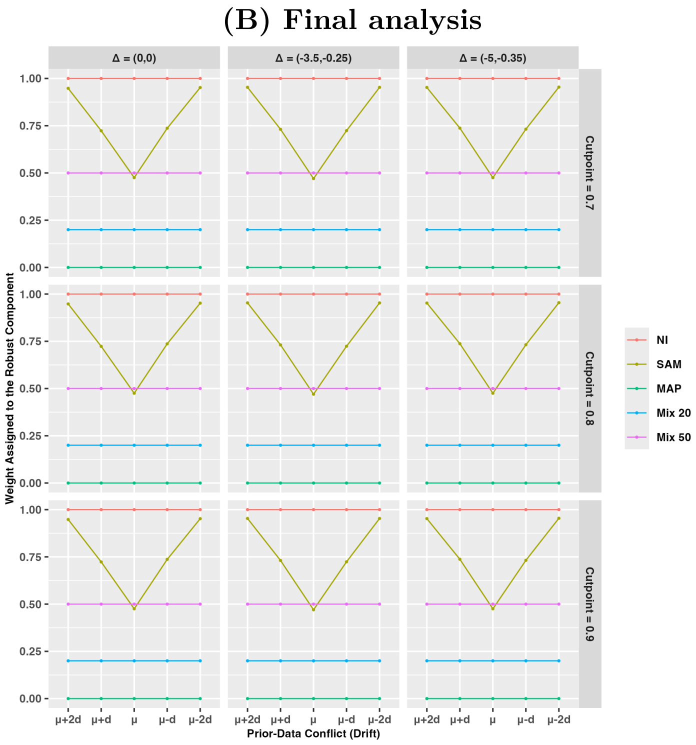


**Figure S7:** Weight assigned to the robust component of the mixture prior for Endpoint 2. Results are shown for the primary analysis using a non-informative prior as the robust component of the mixture prior. Panel (A) shows the weight at the interim analysis, and Panel (B) shows the weight at the final analysis. For the RMAP priors, the weights are fixed by design, whereas for the SAM prior, the weight is adaptively estimated from the observed data. Columns correspond to the three treatment-effect scenarios, and rows correspond to the futility cutoff values ($\gamma = 0.7$, 0.8, and 0.9) used at the interim analysis.

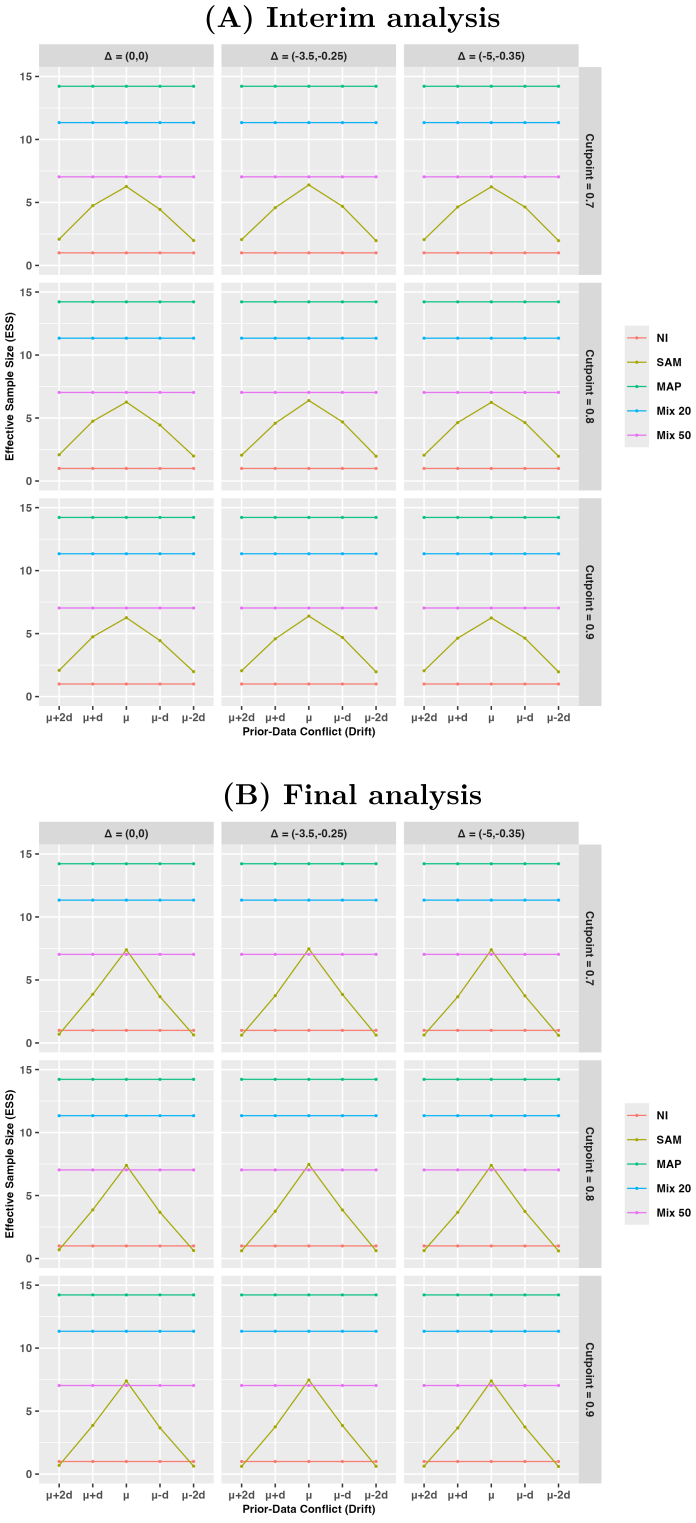


**Figure S8:** Effective sample size (ESS) contributed by the external information for Endpoint 2. Results are shown for the primary analysis using a non-informative prior as the robust component of the mixture prior. Panel (A) shows the ESS at the interim analysis, and Panel (B) shows the ESS at the final analysis. Columns correspond to the three treatment-effect scenarios, and rows correspond to the futility cutoff values ($\gamma = 0.7$, 0.8, and 0.9) used at the interim analysis.

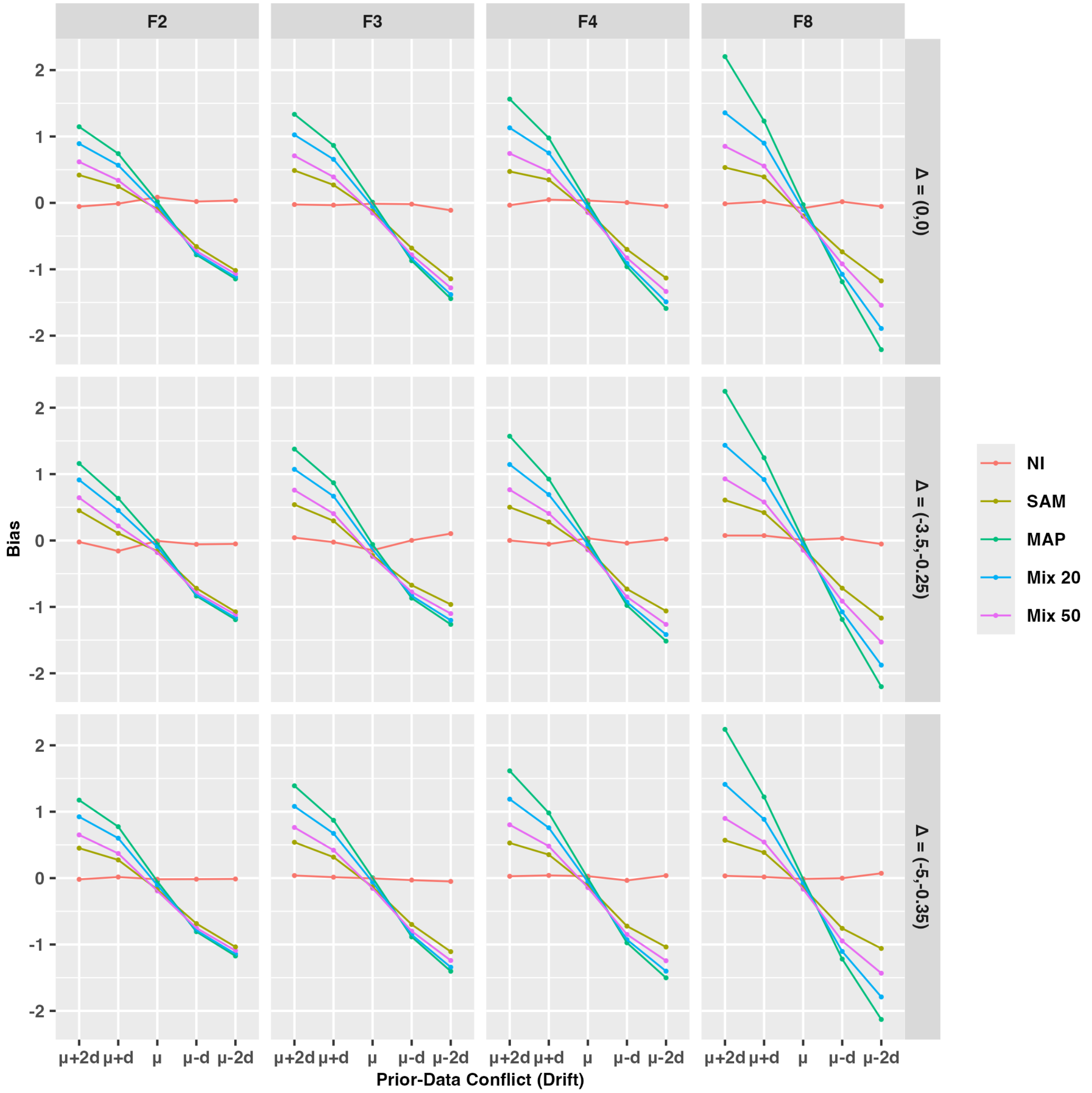


**Figure S9:** Bias for Endpoint 1 as a function of prior–data conflict (drift) and the heterogeneity prior. Results are shown for the sensitivity analysis using a skeptical prior as the robust component of the mixture prior. Columns correspond to the values of $F$ used to specify the prior distribution for the between-study heterogeneity parameter $\tau$, and rows correspond to the three treatment-effect scenarios.

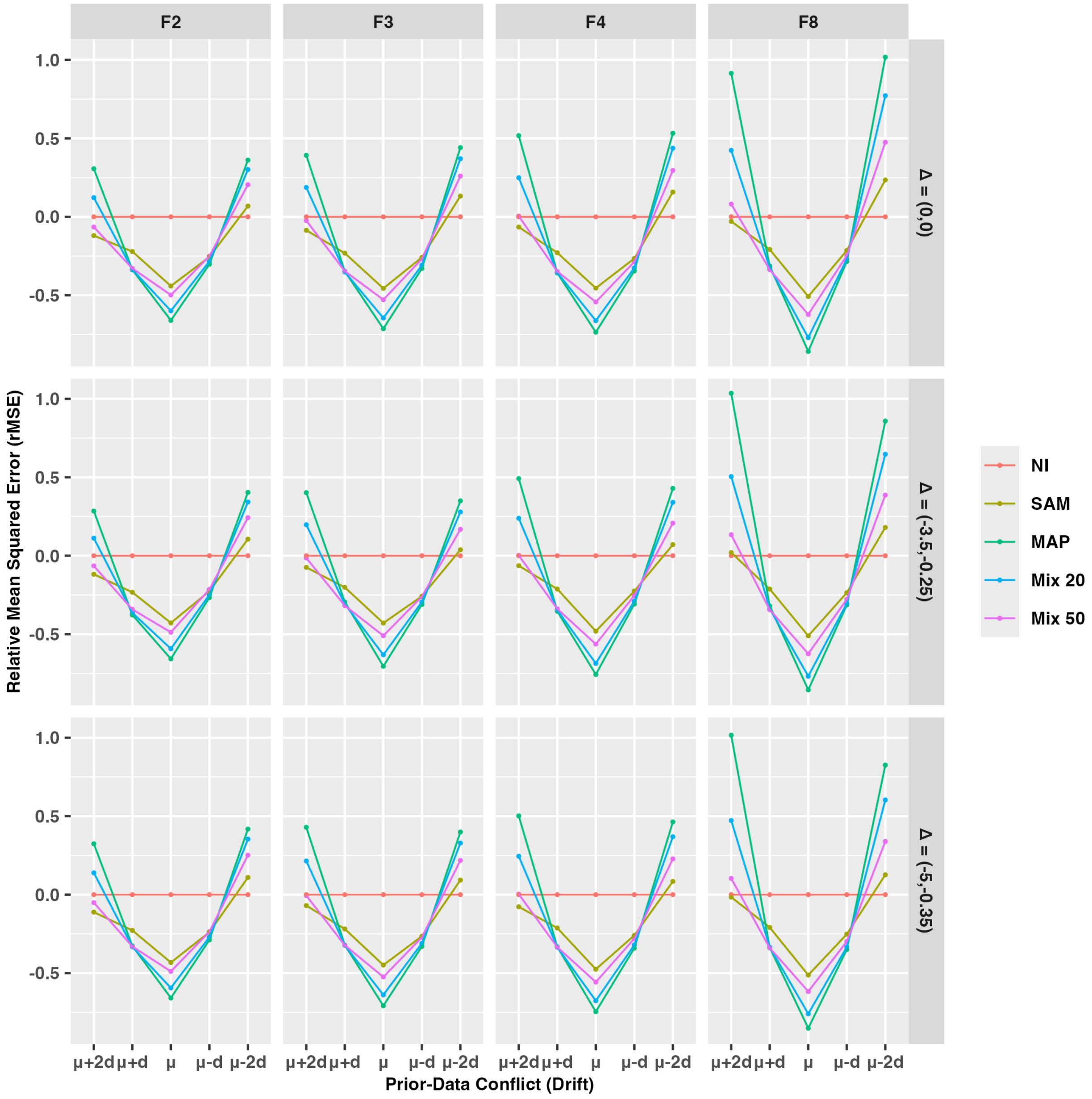


**Figure S10:** Relative mean squared error (rMSE) for Endpoint 1 as a function of prior–data conflict (drift) and treatment-effect scenario. Results are shown for the sensitivity analysis using a skeptical prior as the robust component of the mixture prior. The relative MSE is calculated with respect to the non-informative prior (NI), such that values below zero indicate improved estimation efficiency compared with NI. Columns correspond to the values of $F$ used to specify the prior distribution for the between-study heterogeneity parameter $\tau$, and rows correspond to the three treatment-effect scenarios.

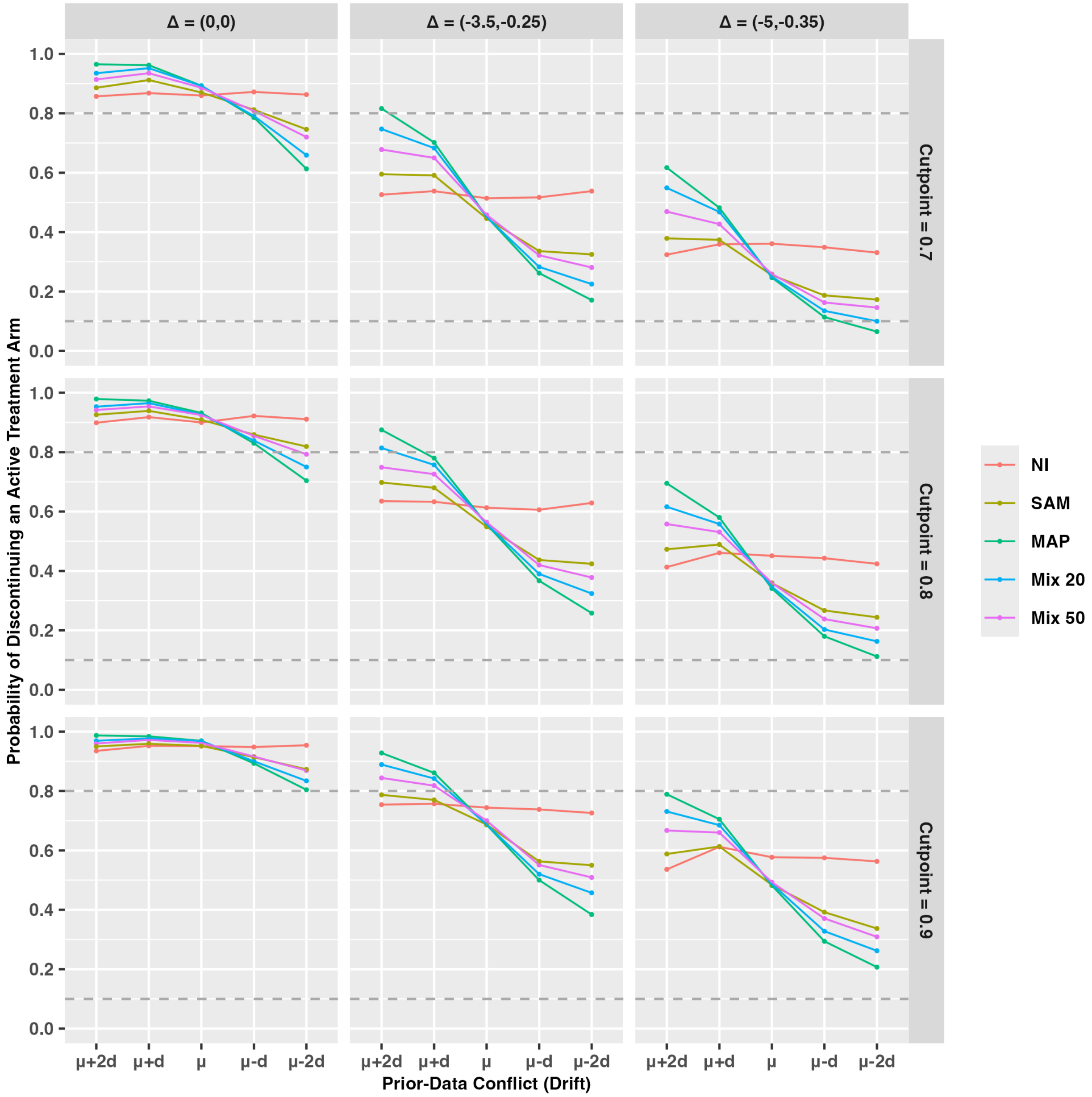


**Figure S11:** Probability of early discontinuation of an active treatment arm at the interim analysis for Endpoint 1 as a function of prior–data conflict (drift) and treatment-effect scenario. Results are shown for the sensitivity analysis using a skeptical prior as the robust component of the mixture prior. Columns correspond to the three treatment-effect scenarios, and rows correspond to the futility cutoff values ($\gamma = 0.7$, 0.8, and 0.9) used at the interim analysis.

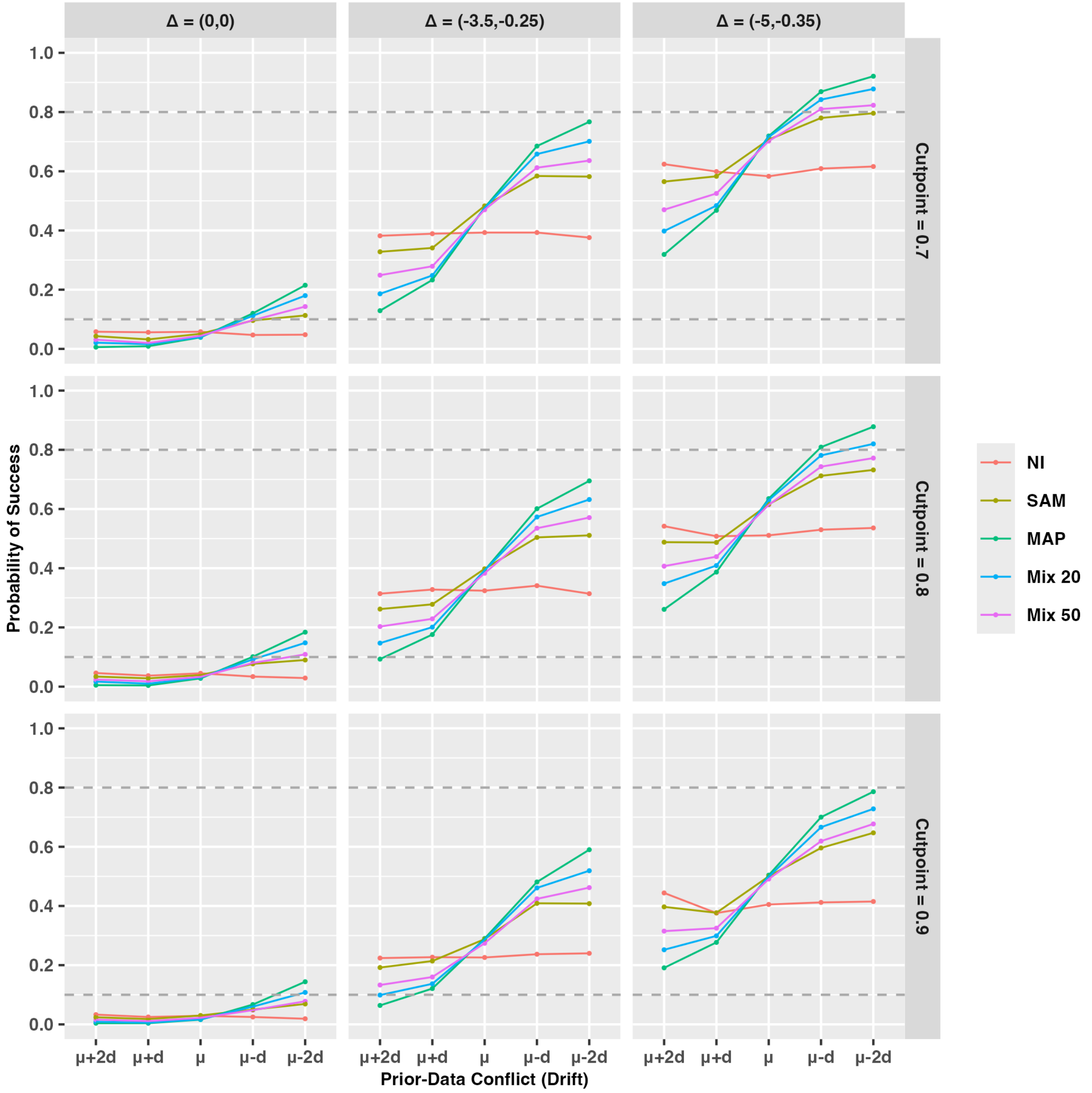


**Figure S12:** Probability of success at the final analysis for Endpoint 1 as a function of prior–data conflict (drift) and treatment-effect scenario. Results are shown for the sensitivity analysis using a skeptical prior as the robust component of the mixture prior. Under the null scenario (i.e., $\Delta = (0, 0)$), the probability of success represents the Type I error rate, whereas under the alternative scenarios it represents statistical power. The horizontal dashed lines indicate the nominal Type I error rate (0.10) and the target power (0.80). Columns correspond to the three treatment-effect scenarios, and rows correspond to the futility cutoff values ($\gamma = 0.7$, 0.8, and 0.9) used at the interim analysis.

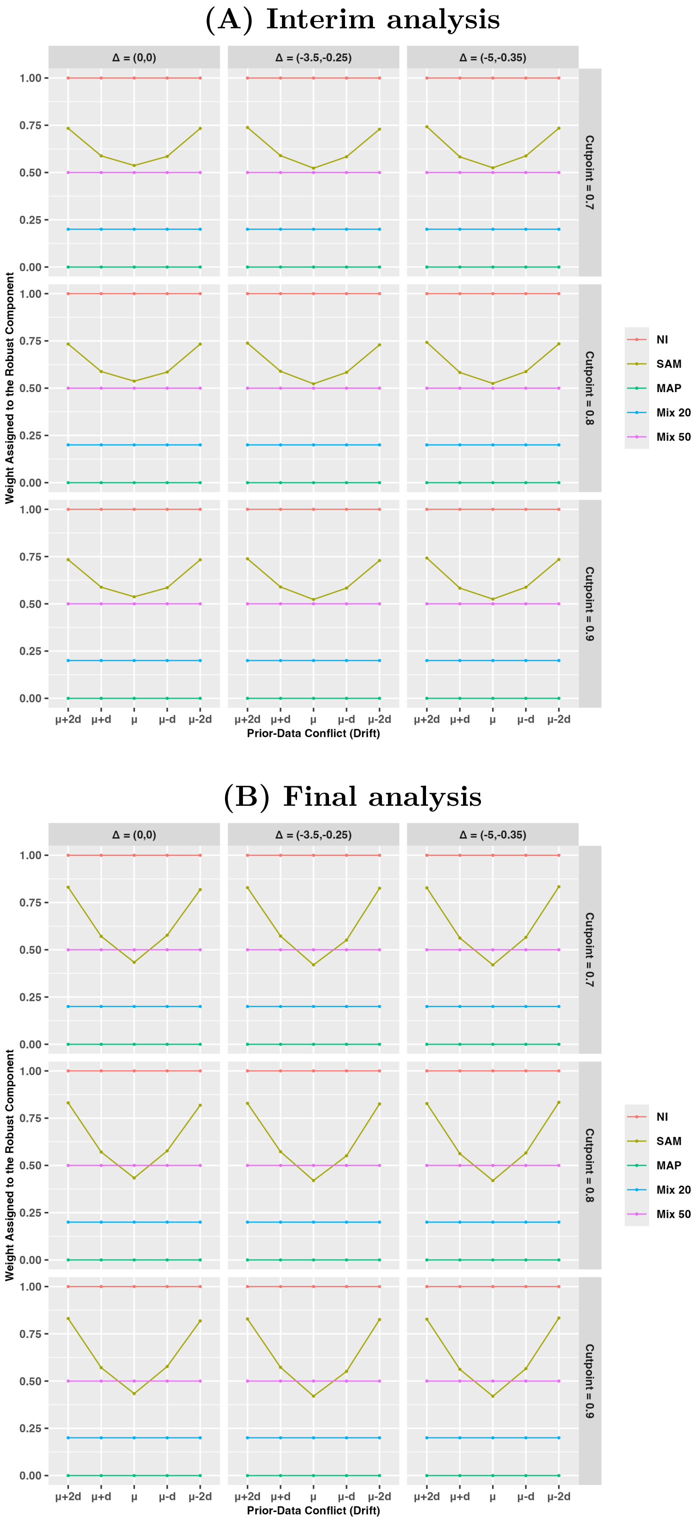


**Figure S13:** Weight assigned to the robust component of the mixture prior for Endpoint 1. Results are shown for the sensitivity analysis using a skeptical prior as the robust component of the mixture prior. Panel (A) shows the weight at the interim analysis, and Panel (B) shows the weight at the final analysis. For the RMAP priors, the weights are fixed by design, whereas for the SAM prior, the weight is adaptively estimated from the observed data. Columns correspond to the three treatment-effect scenarios, and rows correspond to the futility cutoff values ($\gamma = 0.7$, 0.8, and 0.9) used at the interim analysis.

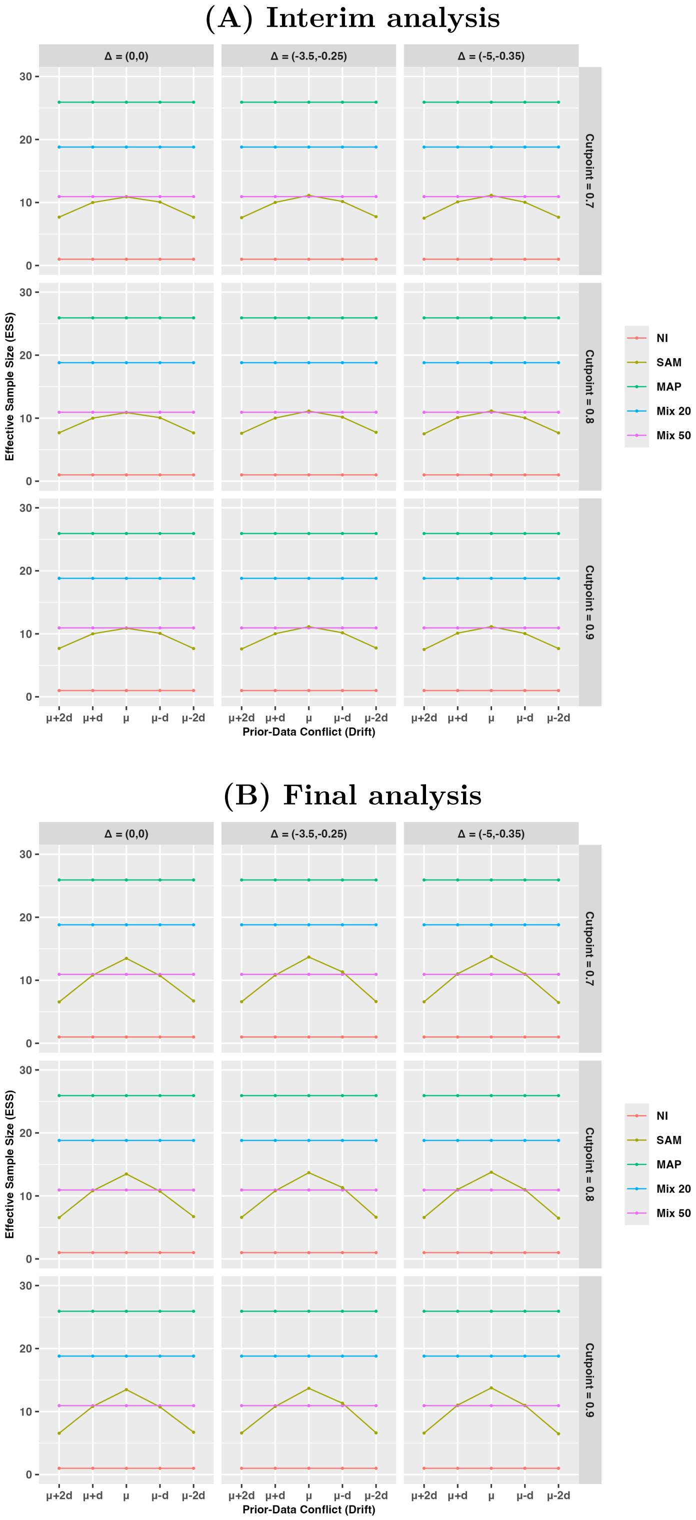


**Figure S14:** Effective sample size (ESS) contributed by the external information for Endpoint 1. Results are shown for the sensitivity analysis using a skeptical prior as the robust component of the mixture prior. Panel (A) shows the ESS at the interim analysis, and Panel (B) shows the ESS at the final analysis. Columns correspond to the three treatment-effect scenarios, and rows correspond to the futility cutoff values ($\gamma = 0.7$, 0.8, and 0.9) used at the interim analysis.
